# Signatures of EMT, immunosuppression and inflammation of primary and recurrent human cutaneous squamous cell carcinoma at single-cell resolution

**DOI:** 10.1101/2022.07.05.22277217

**Authors:** Xin Li, Shuang Zhao, Xiaohui Bian, Lining Zhang, Lixia Lu, Shiyao Pei, Liang Dong, Wensheng Shi, Lingjuan Huang, Xiyuan Zhang, Mingliang Chen, Xiang Chen, Mingzhu Yin

## Abstract

The recurrence of cutaneous squamous cell carcinoma (cSCC) after surgery remains a key factor affecting cSCC outcomes, which is related to the reprogramming of the tumour microenvironment (TME). Herein, we utilized single-cell RNA sequencing (scRNA-seq) to examine the dynamic changes in epithelial cells, T cells, myeloid cells and fibroblasts between primary and recurrent cSCC. We uncovered the immunosuppressed microenvironment in recurrent cSCC, which exhibited a T-cell-excluded and SPP1^+^ TAM-enriched status. In recurrent cSCC, CD8^+^ T cells showed high exhaustion and low inflammatory features, while SPP1^+^ TAMs displayed global protumour characteristics, including decreased phagocytosis and inflammation as well as increased angiogenesis. Furthermore, we found that the subgroups of SPP1^+^ tumour-associated macrophages (TAMs) harboured distinct functions. SPP1^+^ CD209^high^ TAMs showed obvious features of phagocytosis, while SPP1^+^ CD209^low^ TAMs tended to have a high angiogenic ability. A subpopulation of tumour-specific keratinocytes (TSKs) showed significant epithelial–mesenchymal transition (EMT) features in recurrent cSCC, which might be due to their active communication with IL7R**^+^** cancer-associated fibroblasts (CAFs). In addition, we found that MDK could provoke different cell–cell interactions in cSCCs with distinctive staging. In primary cSCC, MDK was highly expressed in fibroblasts and could promote their proliferation and block the migration of tumour cells, while in recurrent cSCC, the high expression of MDK in TSKs promotes their proliferation and metastasis. Overall, our study provides insights into the critical mechanisms of cSCC progression, which might facilitate the development of a powerful system for the prevention and treatment of cSCC recurrence.

## Introduction

cSCC remains the second most common type of nonmelanoma skin cancer after basal cell carcinoma (BCC)(Chang et al., 2022), which originates from epidermal keratinocytes and can develop as an in situ, invasive and finally metastatic form(Ashraf and Alsharedi, 2021; Piipponen et al., 2021). Although most cases of cSCC can be completely eradicated by surgery or ablation, a fraction of these tumours recur, metastasize, and lead to death(Schmults et al., 2013) and are considered high-risk tumours(Martorell-Calatayud et al., 2013). Therefore, there is an urgent need to distinguish high-risk tumours from cSCCs and prevent their progression in the early stage, as this could significantly improve the morbidity and mortality of cSCC. The immunosuppressed phenotype has been observed as an important factor for cSCC recurrence(Yan et al., 2021). Thus, it is worthwhile to characterize the heterogeneous and functional states of cells in the TME of cSCC.

The TME is composed of highly plastic cancer cells, resident and infiltrating host cells, and noncellular tissue components, which constantly interact and collectively determine the progression, metastasis, and therapeutic responses of tumours(Spranger and Gajewski, 2018; Wu et al., 2021a). scRNA-seq technology has achieved an unprecedented revolution into understanding intratumoural transcriptomic heterogeneity in many cancers(Chen et al., 2021) and has revealed the critical cell populations that influence drug resistance and tumour prognosis(Zhang et al., 2021; Patil et al., 2022). A recent study utilized scRNA-seq to dissect the tumour and immune dynamics of cSCC and depicted a global ligand–receptor association of primary human cSCCs and matched normal skin(Ji et al., 2020). However, the heterogeneity between primary and recurrent cSCC and the pathogenesis of cSCC recurrence are not fully understood.

To address this issue, we performed scRNA-seq for 5 patients who were diagnosed with primary or recurrent cSCC to comprehensively analyse the characteristics and alterations of the TME within cSCC. In total, 14,626 single-cell transcriptomes were characterized from tumour tissues and adjacent normal skin (ANS) sites. We identified a subset of TSKs that have remarkable features of EMT in recurrent cSCC. We expanded the knowledge of previous work and revealed that the two subpopulations of SPP1^+^ TAMs display different functions, which were characterized by high phagocytosis (SPP1^+^ CD209^high^ TAMs) and angiogenesis (SPP1^+^ CD209^low^ TAMs) abilities. The primary and recurrent cSCC displayed strikingly distinct compositions of immune cells and fibroblasts, and recurrent cSCC showed a T-cell-excluded and SPP1^+^ TAM-enriched microenvironment. In recurrent cSCC, T cells exhibited high exhaustion and a low inflammatory state, while TAMs displayed low phagocytosis and inflammatory features and a high angiogenic ability. We also identified a group of cancer-associated fibroblasts (CAFs) enriched in recurrent cSCC that closely interact with TSKs. Finally, we found that MDK was upregulated in fibroblasts among primary tumours and potentially promoted the proliferation of fibroblasts, which further blocked the migration of tumour cells from the TME. In recurrent cSCC, MDK might play critical roles in facilitating the proliferation and metastasis of tumour cells, whose expression was upregulated in recurrent tumours and positively correlated with two EMT-associated genes, VIM and TGFB1, based on the IHC staining. Our study emphasizes the potential role of MDK in regulating the alterations of cell–cell crosstalk between primary and recurrent cSCC and may serve as a key factor to mediate cSCC recurrence.

## Results

### A single-cell expression atlas of cSCC ecosystems

To dissect the landscape of the tumour microenvironment (TME) and pathogenesis in primary and recurrent cSCC, we performed scRNA-seq analysis of 9 samples (6 tumour tissues and 3 adjacent normal skin tissues) from patients diagnosed with primary or recurrent cSCC (Fig. 1a, Materials and methods). After quality control and several filtering steps, 14,626 high-quality cells were retained for downstream analysis (Fig. S1a-b).

**Fig. 1.**
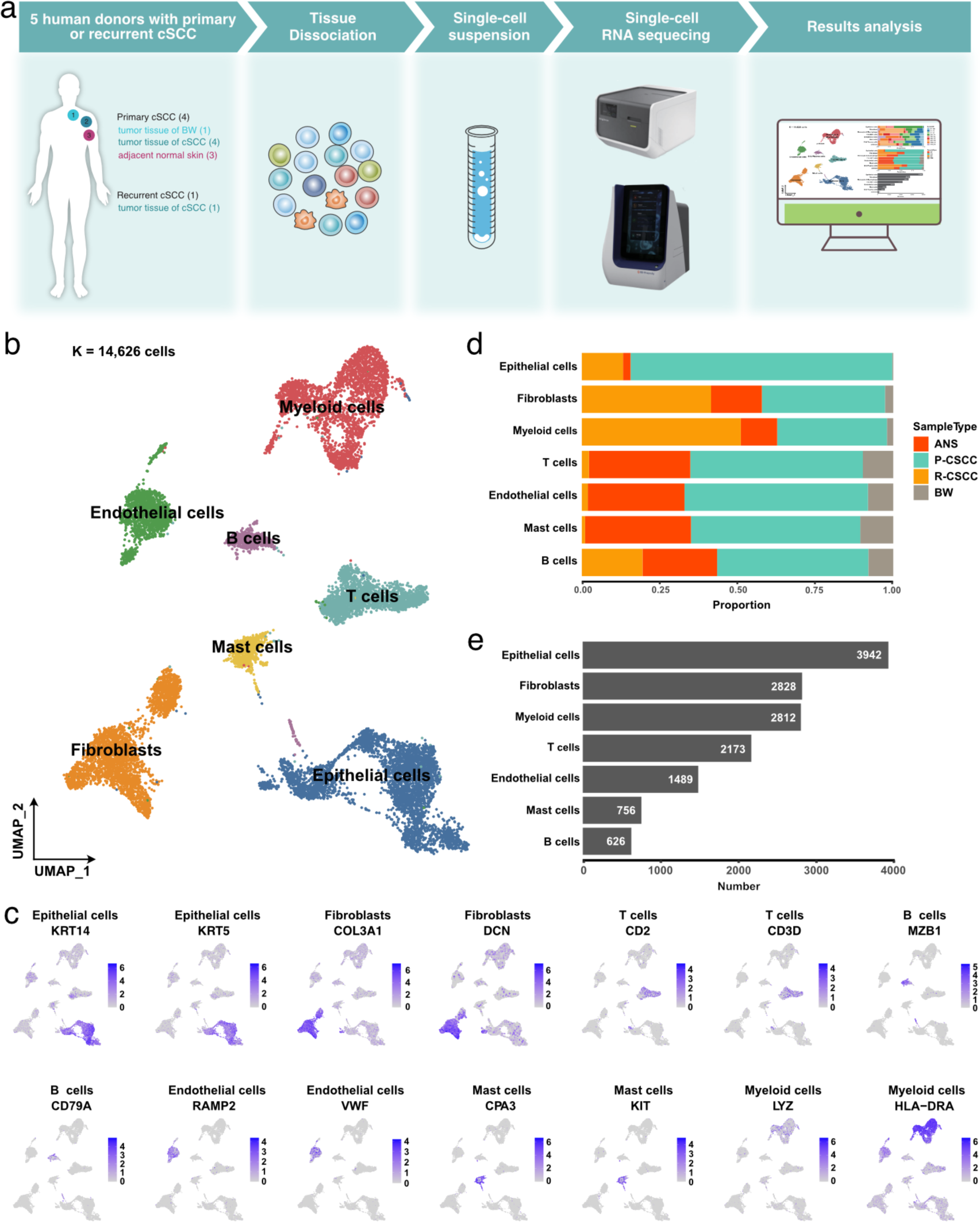
A global overview of TME in cSCC. **a** Schematic graph describing the main workflow and study design. **b** Uniform Manifold Approximation and Projection (UMAP) of major cell populations in our study, different colored dots represent different cell types. **c** The expression level of cell type specific gene markers among UMAP. **d** Sample type fractions relative to the total cell count per cell type. **e** Bar plot shows the cell number of the major cells. ANS, adjacent normal skin. BW, Bowen disease. P-cSCC: primary cutaneous squamous cell carcinoma. R-cSCC: recurrent cutaneous squamous cell carcinoma.

All scRNA-seq data were merged together, and gene expression normalization and scaling, dimension reduction, batch correction, and cell clustering were performed to identify coarse cell types. Seven major cell types were detected based on the gene expression of canonical cell markers, including epithelial cells, fibroblasts, myeloid cells, T cells, endothelial cells, mast cells, and B cells (Fig. 1b-c, Materials and methods). The proportions of these major cell types varied greatly among different samples (Fig. S1c), suggesting the heterogeneous character of the TME in cSCC. For most cell types, tumour tissue of primary cSCC contains the highest cell abundance, tumour tissue of recurrent cSCC contains a relatively high proportion of fibroblasts and B cells, while tumour tissue of Bowen disease (BW) contains the lowest cell proportion (Fig. 1d). Overall, epithelial cells, fibroblasts and myeloid cells were the main components of the microenvironment (Fig. 1e).

### Tumour-specific keratinocytes show obvious EMT characteristics in recurrent cSCC

Overall, 3,942 epithelial cells were further reclassified into 4 clusters. In this process, we removed cell clusters that also expressed the gene markers of other cells, leaving 2,371 cells (Materials and methods, Fig. S2). Cells in each of the four clusters expressed known representative genes (Fig. 2a, Fig. S3a): (1) basal (COL17A1+), (2) cycling (MKI67+, TOP2A+), (3) differentiating (KRT1+), and (4) TSK (MMP10+, PTHLH+) cells(Ji et al., 2020).

**Fig. 2.**
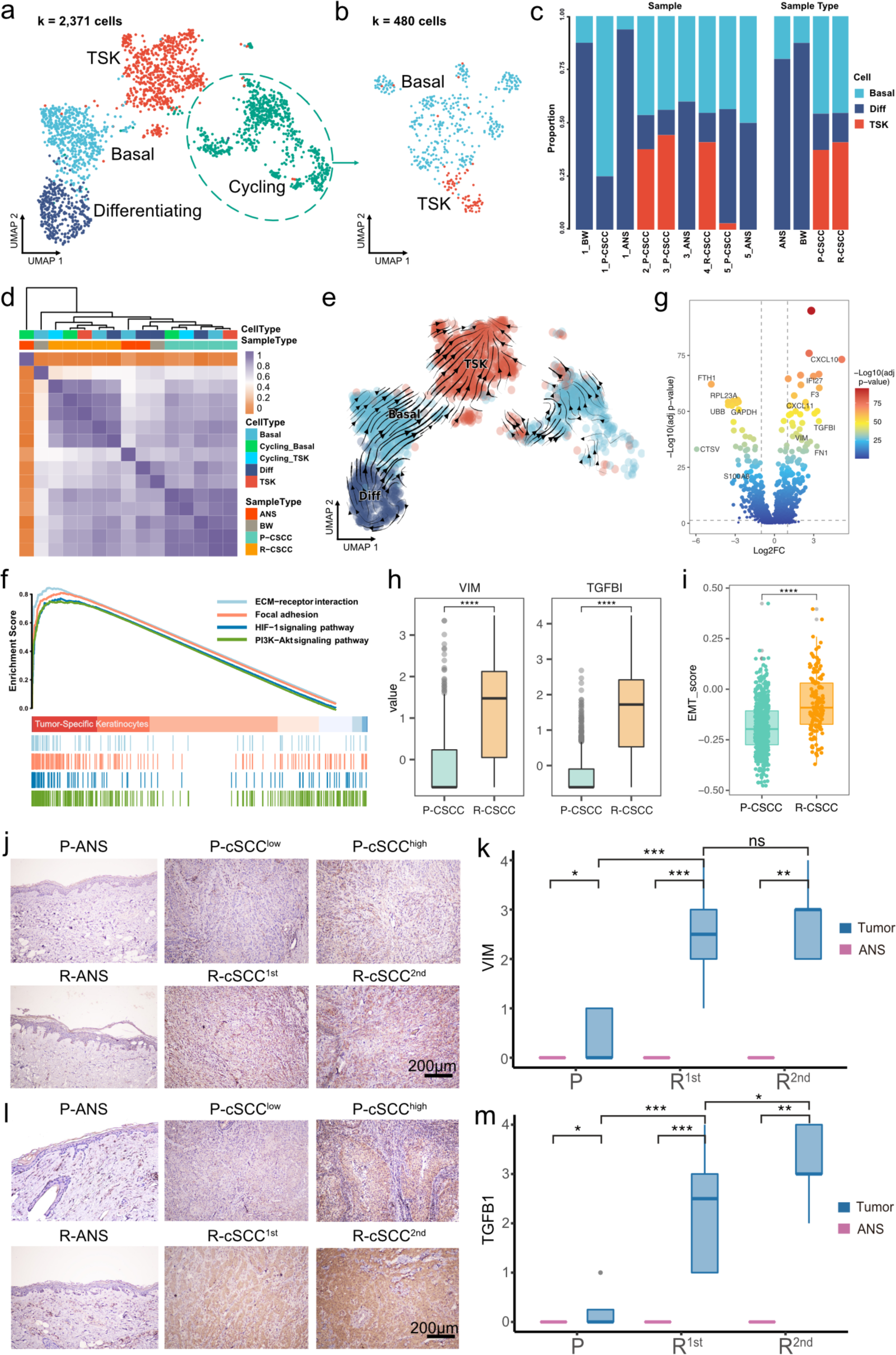
Re-clustering and functional analysis of epithelial cells. **a** UMAP plot of sub- groups of epithelial cells. **b** UMAP plot of cycling cells. **c** The proportion of different cell types among samples and tissues. **d** Heatmap depicting pairwise correlations between different cell types. **e** Cell transition potential of basal, differentiating cells and TSKs determined by RNA velocity analysis. **f** Enriched pathways of TSKs by GSEA analysis. **g** Volcano plot showing the differential expressed genes between primary and recurrent cSCC. **h** Gene expression level of VIM and TGFB1 across primary and recurrent cSCC. **i** MET signature score of TSKs across primary and recurrent cSCC. **j** The IHC staining of VIM in representative samples. **k** The IHC score of VIM in primary, the first and second recurrence of cSCC, different tissue sites were marked by different color. **l** The IHC staining of TGFB1 in representative samples. **m** The IHC score of TGFB1 in primary, the first and second recurrence of cSCC, different tissue sites were marked by different color. Wilcoxon signed-rank test, \*\*\*\**p* < 0.0001.

Furthermore, the cycling cells were grouped into cycling TSKs and basal cells based on the expression of known markers (Fig. 2b, Fig. S3b). Overall, the ANS and BW had a high proportion of differentiating cells, while primary and recurrent tumour tissues had a relatively high proportion of TSKs (Fig. 2c). Correlation analysis showed that epithelial cells from the same sample type were prone to cluster together (Fig. 2d), demonstrating that epithelial cells are highly heterogeneous relative to sample site origin. RNA velocity analysis revealed that basal and differentiating cells tended to differentiate into TSKs (Fig. 2e). Functional enrichment analysis results showed that TSK cells are associated with the PI3K-Akt signalling, ECM-receptor interaction, focal adhesion, and HIF-1 signalling pathways (Fig. 2f), which have been demonstrated to be associated with tumour progression or resistance to cancer therapies(Fresno Vara et al., 2004; Eke and Cordes, 2015; Masoud and Li, 2015; Bao et al., 2019; He et al., 2021). We further evaluated the activity score of functional terms associated with “hallmarks of cancer”. TSK cells showed the highest scores in most hallmarks, demonstrating they have cancer characteristics, such as “Self Sufficiency in Growth Signals”, “Insensitivity to Antigrowth Signals”, and “Evading Apoptosis” (Fig. S3c). Furthermore, we identified differentially expressed genes in TSK cells between primary and recurrent cSCC and found that two EMT-related genes, VIM and TGFB1, were upregulated in recurrent cSCC (Fig. 2g-h). Moreover, we calculated the EMT signature score of TSKs and found that TSKs possessed significantly higher EMT scores in recurrent cSCC (Fig. 2i). Next, to validate the expression of VIM and TGFB1 in cSCC, we performed IHC staining in a clinical cohort that comprised 16 patients (all of them had cancer progression events, from primary tumours to cSCC with single or multiple recurrences, Table S1). The results showed that both VIM and TGFB1 were significantly highly expressed in recurrent cSCC (Fig. 2j-m). This implies that the recurrence of cSCC is associated with EMT.

### Recurrent cSCC displays a T-cell-excluded microenvironment

Subclustering of T cells identified five subpopulations: (1) effector CD4^+^ T cells (CD4^+^, CCR7^-^), (2) Tregs (CD4^+^, IL2RA^+^, FOXP3^+^, CTLA4^+^), (3) naïve CD4^+^ T cells (CD4^+^, CCR7^+^, SELL^+^), (4) effector CD8^+^ T cells (CD8^+^, GZMA^+^), and (5) CD8^+^ cytotoxic cells (CD8^+^, IFNG^+^, GZMA^+^) (Fig. 3a-b). RNA velocity analysis displayed bidirectional flows between Tregs and effector CD4^+^ T cells, effector CD8^+^ T cells and CD8^+^ cytotoxic cells, while the differentiation from naïve CD4^+^ T cells to Tregs seemed to be irreversible (Fig. S4a). This finding indicated that Tregs, effector CD4^+^ T cells, effector CD8^+^ T cells and CD8^+^ cytotoxic cells in cSCC were prone to becoming intermediate and plastic and had the potential to differentiate into other cells.

**Fig. 3.**
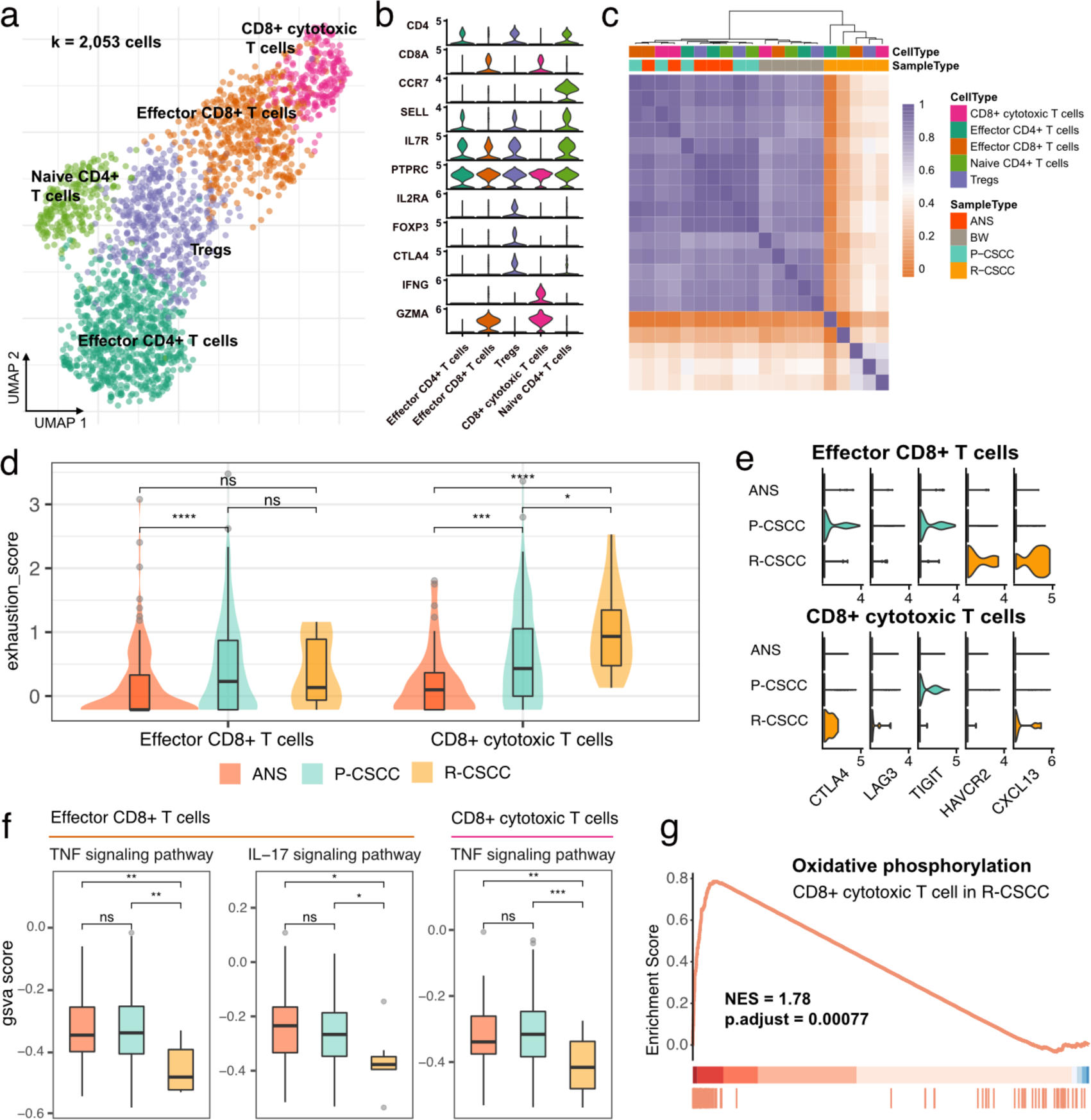
T cell annotation and functional characterization. **a** UMAP visualization of T cells, different colors represent distinct sub-populations. **b** Violin plot showing the representative markers of T cell lineage. **c** Heatmap showing the pairwise correlations between T cells. **d** The distribution of exhaustion score of effector CD8+ T cells and CD8+ cytotoxic T cells among ANS, primary and recurrent cSCC. **e** The expression level of immune inhibitors among different sample and cell types. **f** The GSVA score of inflammatory pathways in effector CD8+ T cells and CD8+ cytotoxic T cells among different sample types. **g** Significantly identified pathway that enrich in recurrent versus primary cSCC. Wilcoxon signed-rank test, \**p* < 0.05, \*\**p* < 0.01, \*\*\**p* < 0.001.

Both CD4^+^ and CD8^+^ T cells were enriched in primary cSCC, while recurrent cSCC sites showed the lowest T-cell infiltration (Fig. S4b-c), indicating that recurrent cSCC tends to be a T-cell desert tumour. The above-described T cells from different populations were further clustered based on their gene expression level (Materials and methods). The results showed that T cells from ANS and primary cSCC tended to cluster together, and T cells from recurrent cSCC had a distinct pattern relative to other sites of origin (Fig. 3c). Next, we evaluated the exhaustion score of effector CD8+ T cells and CD8+ cytotoxic T cells and compared it among ANS, primary cSCC and recurrent cSCC (Materials and methods). Compared with ANS, both effector CD8+ T cells and CD8+ cytotoxic T cells showed increased exhaustion scores in primary cSCC, while CD8+ cytotoxic T cells had increased exhaustion scores in recurrent cSCC when compared with ANS and primary cSCC (Fig. 3d). Specifically, in primary cSCC, CTLA4 and TIGIT displayed high expression levels in effector CD8+ T cells, and TIGIT showed high expression levels in CD8+ cytotoxic T cells; in recurrent cSCC, effector CD8+ T cells exhibited high expression levels of HAVCR2 (TIM3) and CXCL13, while CD8+ cytotoxic T cells displayed high expression levels of CTLA4 and CXCL13 and relatively higher expression levels of LAG3 (Fig. 3e). This observation indicated that the exhaustion of CD8+ T cells in primary and recurrent cSCC was related to different inhibitors.

Furthermore, to gain deeper insight into the inflammatory changes among the ANS and primary and recurrent cSCC, we performed an inflammatory score evaluation, focusing on 4 pathways (Materials and methods). For effector CD8+ T cells, primary cSCC exhibited activity scores similar to those of ANS, while recurrent cSCC showed significantly decreased inflammatory scores in the TNF signalling pathway and IL-17 signalling pathway (Fig. 3f, Fig. S4d). For CD8+ cytotoxic T cells, except for the IL- 17 signalling pathway, primary cSCC displayed an inflammatory score similar to that of ANS, while the activity scores for the TNF signalling pathway and NOD-like receptor signalling pathway showed significantly decreased levels in recurrent cSCC (Fig. 3f, Fig. S4e). In addition, in CD8+ cytotoxic T cells, GSEA showed that the “oxidative phosphorylation” process was upregulated in recurrent cSCC compared to primary cSCC (Fig. 3g, NES = 1.78, *p* < 0.01), demonstrating that cells were in a metabolically active state. Together, our analysis revealed that recurrent cSCC exhibited low infiltration, a distinct pattern and a decreased inflammatory score of T cells, which may be a prominent reason for cSCC recurrence.

### SPP1^+^ CD209^high/low^ TAMs are characterized by several protumour phenotypes in recurrent cSCC

Myeloid cells were some of the most abundant cells in the TME of cSCC (Fig. 1e) and have been demonstrated to participate in tumour progression and metastasis(Kaczanowska et al., 2021). Subsequently, 2,693 myeloid cells were clustered and annotated into 10 subpopulations (Fig. 4a). In total, 4 populations were designated TAMs, which had various features: (1) SPP1^+^ CD209^high^ TAMs (SPP1^+^, CD209^high^, CD163^+^, MRC1^+^, CCL18^+^), (2) SPP1^+^ CD209^low^ TAMs (SPP1^+^, CD209^low^, CD163^+^, MRC1^+^, CCL18^+^), (3) CXCL10^+^ TAMs (CD163^+^, MRC1^+^, CXCL10^+^), and (4) cycling TAMs (CD209^+^, CD163^+^, MRC1^+^, TOP2A^+^, MKI67^+^). In addition, one cluster was identified as monocytes (VEGFA^+^, VCAN^+^, FCN1^+^), and three subpopulations were characterized as DCs: (1) CD14^+^ DCs (CD1A^-^, CD14^+^, CD1C^+^), (2) CLEC9A^+^ DCs (CLEC9A^+^, CD1C^+^), and (3) CD1a^+^ CD1c^+^ DCs (CLEC10A^+^, CD1A^+^, CD1C^+^, CD14^-^). In addition, we divided MDSCs into two subgroups: (1) CXCL9-11^+^ MDSCs (CXCL9^+^, CXCL10^+^, CXCL11^+^, IL1B^+^, S100A8^+^, S100A9^+^) and (2) CXCL1-3^+^ MDSCs (CXCL1^+^, CXCL2^+^, CXCL3^+^, IL1B^+^, S100A8^+^, S100A9^+^).

**Fig. 4.**
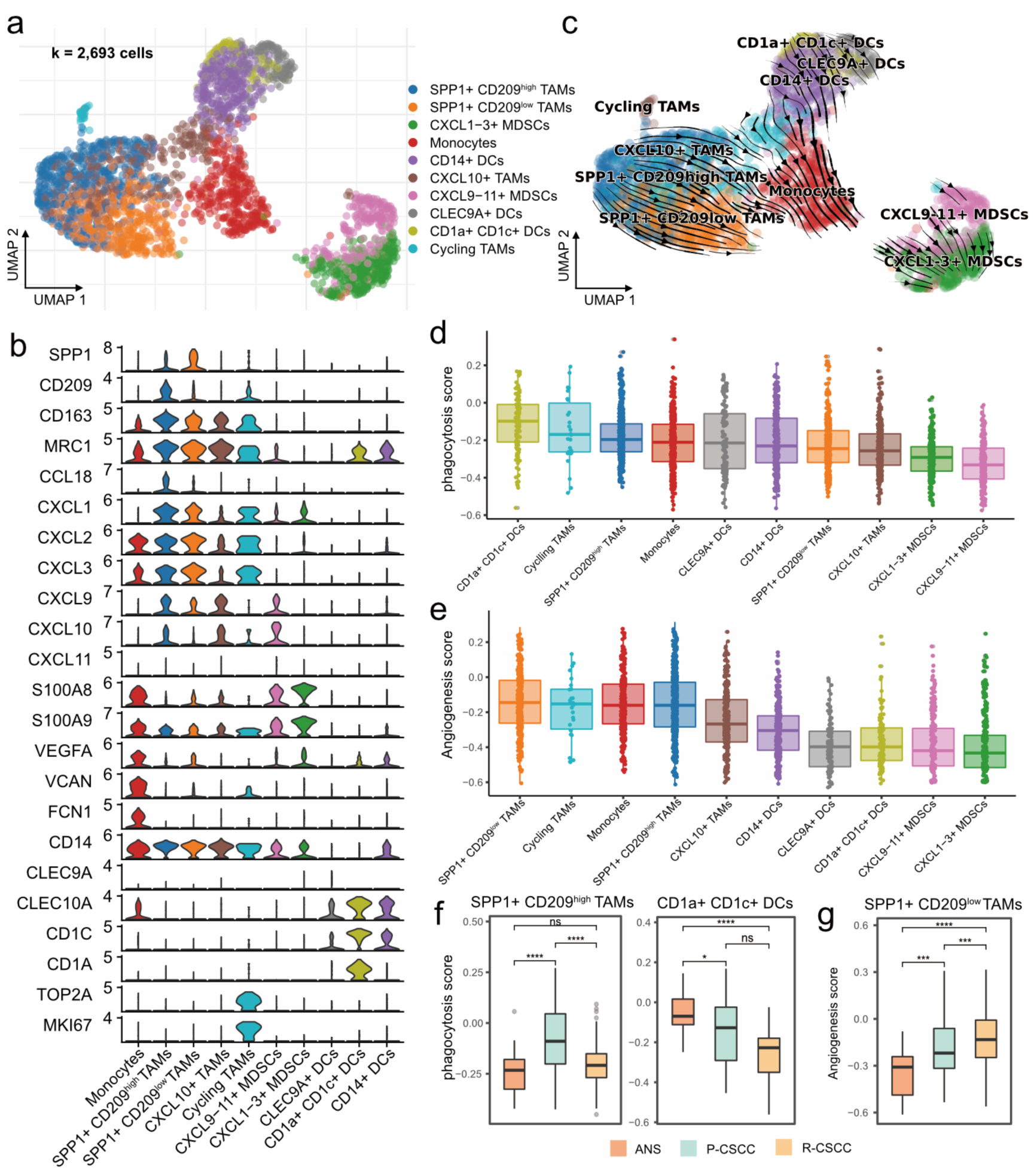
Components and phenotypes of myeloid cells in cSCC. **a** UMAP plot of myeloid cells. **b** Violin plot of the maker genes in myeloid sub-populations. **c** UMAP plot showing the RNA velocity of myeloid cells. **d** Distribution of phagocytosis score in each cell type, ranking by the median value. **e** Distribution of angiogenesis in each cell type, ranking by the median value. **f** The GSVA score of phagocytosis score in SPP1^+^ CD209^high^ TAMs and CD1a^+^ CD1c^+^ DCs among different sample types. **g** The GSVA score of angiogenesis in SPP1^+^ CD209^low^ TAMs among different sample types. Wilcoxon signed-rank test, \**p* < 0.05, \*\*\**p* < 0.001, \*\*\*\**p* < 0.0001.

The expression of each cell-specific gene marker is presented in Fig. 4b and Fig. S5. Notably, for the two groups of MDSCs identified in our study, CXCL9-11^+^ MDSCs were enriched in recurrent cSCC, while CXCL1-3^+^ MDSCs were more abundant in primary cSCC (Fig. S6a-b). Compared with CXCL1-3^+^ MDSCs, CXCL9-11^+^ MDSCs were enriched in the pathways of “oxidative phosphorylation” and “antigen processing and presentation” according to GSEA (Fig. S6c).

RNA velocity analysis showed the transition directions from TAMs and DCs to monocytes, and MDSCs were located at the end of the differentiation trajectory (Fig. 4c). Furthermore, we confirmed that the potential origins of monocytes were M2 macrophages and DCs using CytoTRACE(Gulati et al., 2020) (Fig. S6d). Recent studies reported that *Bordetella pertussis* adenylate cyclase toxin could inhibit the differentiation of infiltrating monocytes into macrophages and dendritic cells by activating cMAP signalling and could provoke the dedifferentiation of macrophages to monocyte-like cells (Ahmad and Sebo, 2020). We thus examined the cMAP activity score of myeloid cells over time. Interestingly, there was a significantly negative correlation between latent time and cMAP score (R = 0.32, *p* < 0.00001). The cMAP score displayed a decreased pattern when macrophages and DCs transformed into monocytes, and we found no significant reduction pattern in cMAP score for cells that originated from monocytes (Fig. S6e-f). These observations suggest that the transmission of monocytes from macrophages and DCs may be related to the cMAP signalling pathway and that the transmission direction may be related to the malignancy level of cSCC.

Recently, a subtype of TAMs with SPP1^+^ characteristics was reported to carry angiogenesis-related properties and was linked to tumour metastasis(Cheng et al., 2021; Liu et al., 2022; Qi et al., 2022). In our study, SPP1^+^ CD209^high^ and SPP1^+^ CD209^low^ TAMs were identified as two subpopulations of SPP1^+^ TAMs, which showed an elevated proportion in recurrent cSCC (Fig. S6a-b). We examined the angiogenic and phagocytotic properties of TAMs and found that they showed different characteristics. SPP1^+^ CD209^high^ TAMs exhibited a higher phagocytosis score than SPP1^+^ CD209^low^ TAMs, while SPP1^+^ CD209^low^ TAMs harboured the highest angiogenesis score (Fig. 4d-e). In addition, CD1a^+^ CD1c^+^ DCs carried the highest phagocytosis score (Fig. 4d). Not surprisingly, the phagocytosis score of SPP1^+^ CD209^high^ TAMs, CD1a^+^ CD1c^+^ DCs and cycling TAMs decreased in recurrent cSCC compared with primary cSCC (Fig. 4f, Fig. S6 g). In addition, SPP1^+^ CD209^low^ TAMs displayed continuously significantly increased angiogenesis scores in primary and recurrent cSCC (Fig. 4g, Fig. S6h). We next evaluated the inflammatory pathway scores of these myeloid cells. Overall, TAMs and monocytes tended to have higher inflammatory scores (Fig. S7a-d). For the TNF, NF-kappa B and NOD-like receptor signalling pathways, SPP1^+^ CD209^high^ TAMs had elevated scores in primary cSCC and displayed decreased scores in recurrent cSCC (Fig. S7e, g-h). For the NF-kappa B and IL-17 signalling pathways, monocytes showed lower scores in primary and recurrent cSCC than in ANS (Fig. S7e-f). Taken together, our results revealed that SPP1^+^ TAMs displayed lower phagocytosis and inflammation scores and higher angiogenesis scores in recurrent cSCC, which potentially reflects the cSCC recurrence mechanism.

### Recurrent cSCC-enriched IL7R^+^ CAFs have close crosstalk with tumour-specific keratinocytes

In our study, 2,828 fibroblasts were detected and classified into 3 subpopulations based on the expression of marker genes, including (1) mCAFs (RGS5^+^, DCN^+^, COL6A2^+^, COL1A1^+^, COL1A2^+^), (2) iCAFs (RGS5^-^, DCN^+^, COL6A2^+^, COL1A1^+^, COL1A2^+^), and (3) IL7R**^+^** CAFs (IL7R^+^, IL1B^+^, IL6^+^, CXCL1^+^, CXCL3^+^, CXCL5^+^, CXCL6^+^, CXCL8^+^, CXCL13^+^, CXCL14^+^, DCN^+^, COL6A2^+^, COL1A1^+^, COL1A2^+^) (Fig. 5a-b).

**Fig. 5.**
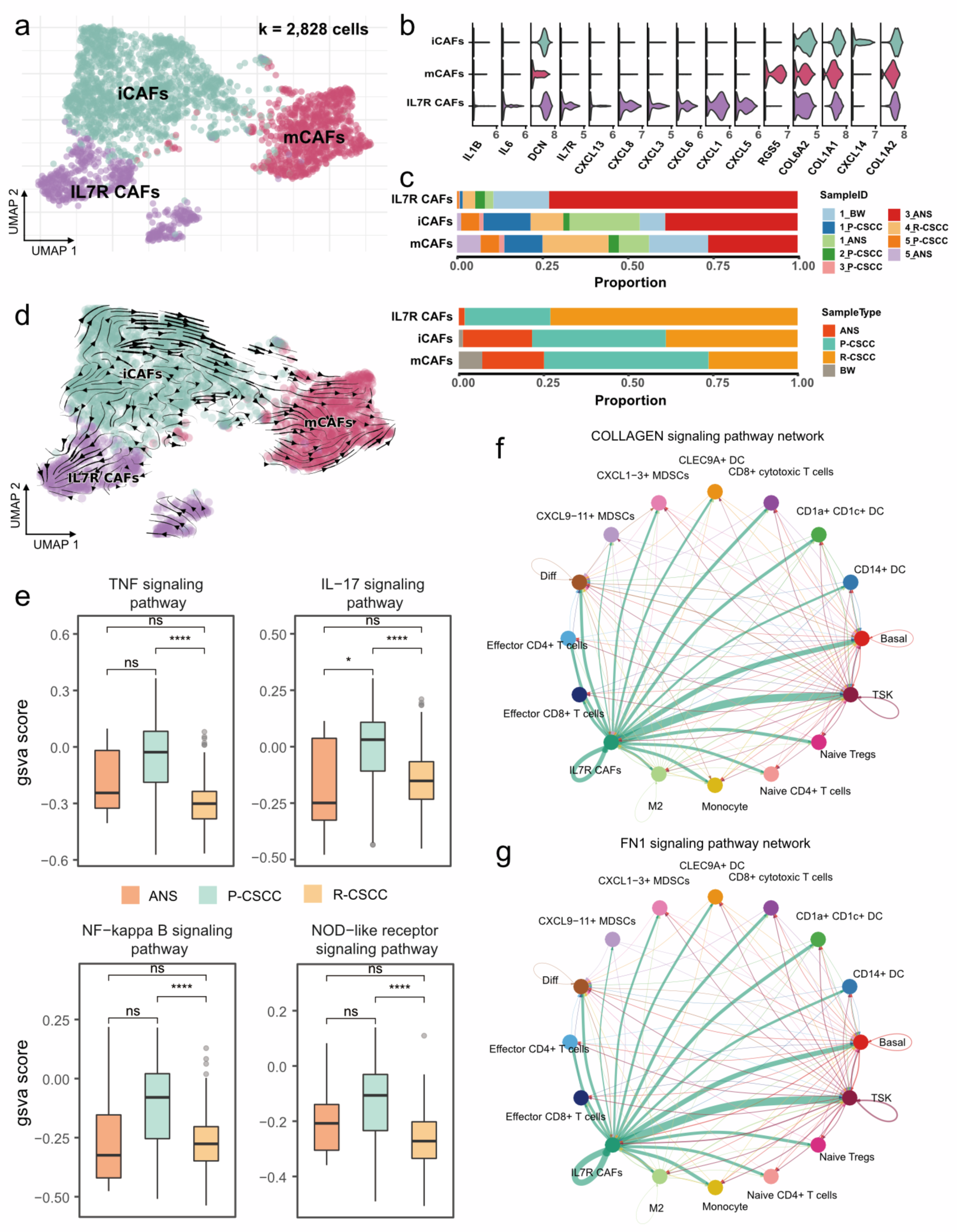
Assessing the functional states of fibroblasts in cSCC. **a** UMAP plot showing the sub-populations of fibroblasts. **b** Violin plot showing the high expression of gene markers in fibroblasts. **c** The proportion of fibroblasts relative to sample ID and sample type. **d** RNA velocity plot showing cell transition directions among fibroblasts. **e** The GSVA score of inflammatory pathways in fibroblasts among different sample types. **f** Cell-cell interactions in COLLAGEN signaling pathway. **g** Cell-cell interactions in FN1signaling pathway. Wilcoxon signed-rank test, \*\*\*\**p* < 0.0001.

Notably, IL7R**^+^** CAFs were more abundant in recurrent cSCC, while primary cSCC had a relatively high proportion of mCAFs (Fig. 5c). RNA velocity analysis revealed the transition direction from iCAFs to mCAFs, as well as iCAFs to IL7R**^+^** CAFs (Fig. 5d), indicating that iCAFs could differentiate into mCAFs and IL7R**^+^** CAFs, which may be related to primary and recurrent cSCC, respectively.

Next, we calculated the inflammatory score of IL7R**^+^**CAFs and compared it between ANS, primary and recurrent cSCC. We found that primary cSCC showed a slight increase in the inflammatory score in all four investigated pathways, while the inflammatory score was significantly decreased in recurrent cSCC (Fig. 5e). This indicates that IL7R**^+^** CAFs tend to display a low inflammatory feature when cSCC recurs. To gain deeper insight into the biological function of IL7R**^+^** CAFs, we performed cell–cell communication analysis using CellChat(Jin et al., 2021). Interestingly, IL7R**^+^** CAFs expressed high levels of valid ligands related to EMT, such as collagens [encoded by COL1A1, COL1A2, COL4A1, COL6A1, COL6A2, COL6A3], Fibronectin 1 [encoded by FN1], Tenascin C [encoded by TNC], and Thy-1 [encoded by THY1] (https://www.gsea-msigdb.org/gsea/msigdb/cards/HALLMARK_EPITHELIAL_MESENCHYMAL_TRANSITION.html), whose receptors were expressed by a wide range of cells, which subsequently induced typical cell–cell interactions (Fig. 5f-g, Fig. S8a-f). Notably, IL7R**^+^**CAFs showed the strongest interaction with TSKs in the Collagen signalling pathway, FN1 signalling pathway and TEAD signalling pathway. For the receptors that are expressed in TSKs, integrins [encoded by ITGA1, ITGA2, ITGA3, ITGA4, ITGA5, ITGA8, ITGAV, ITGAX, ITGAM, ITGB1, and ITGB2] can recognize multiple ligands, including collagens, Fibronectin 1, Tenascin C and Thy-1, which causes downstream crosstalk between TSKs and IL7R**^+^** CAFs. Integrin signalling is known to have a profound effect on tumour cells, including proliferation, migration and survival(Desgrosellier and Cheresh, 2010). ITGA3 was proven to participate in promoting endothelial cell motility and angiogenesis during the early stages of neovascularization(Fukushi et al., 2004); ITGA8 has been demonstrated to regulate the recruitment of mesenchymal cells into epithelial structures and promote cell survival(Lu et al., 2002; Farias et al., 2005). Therefore, IL7R**^+^** CAFs might promote the EMT of TSKs by cell–cell communication through multiple signalling pathways in recurrent cSCC.

### Cell–cell interaction analysis identifies the role of the MDK-dependent pathway in recurrent cSCC

Early evidence suggested differences in the composition and functional status of the TME between primary and recurrent cSCC; thus, we hypothesized that primary and recurrent cSCC display distinct subcellular interaction relationships within the TME, which may have a profound influence on the tumour phenotype. To test this hypothesis, we performed cell–cell communication analysis in primary and recurrent cSCC separately. The results showed that primary cSCC had many more cell interaction events than recurrent cSCC (6696 vs. 1196), which may have been due to the small cell number in recurrent cSCC (8217 vs. 3341). Notably, we observed that iCAFs and IL7R**^+^**CAFs had more interactions with other cells in primary cSCC, whereas TSKs, iCAFs, mCAFs and IL7R**^+^**CAFs were dominant cells that communicated with other cells in recurrent cSCC (Fig. 6a). In addition, we calculated the incoming and outgoing interaction strengths of cells and found that TSKs play an important role within the TME in recurrent cSCC compared with primary cSCC (Fig. S9a).

**Fig. 6.**
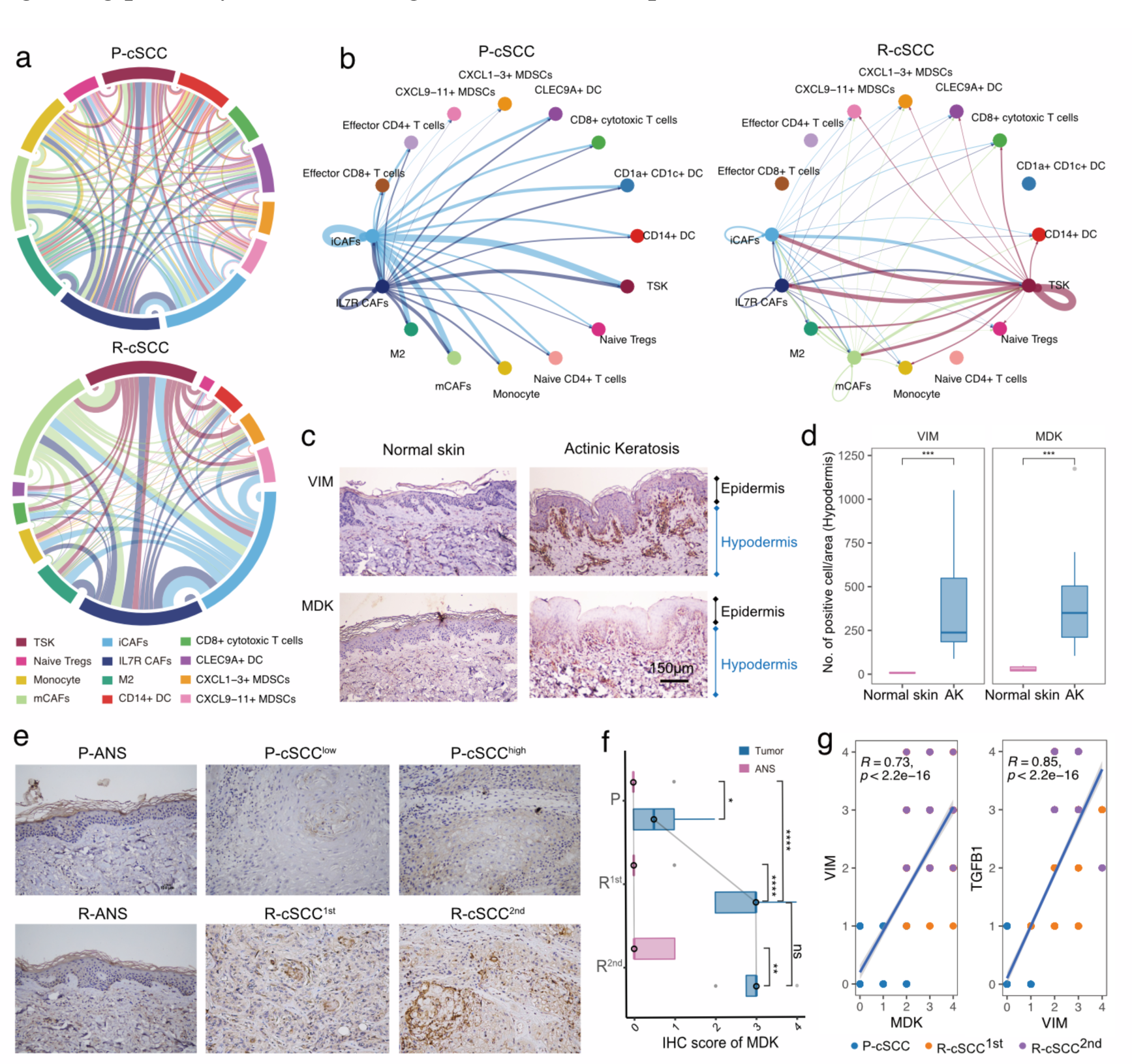
Cell-cell interactions of the primary and recurrent cSCC. **a** Dependency wheel illustrating the interaction relationship between cells within the TME of primary and recurrent cSCC, the link size represents the interaction number. **b** Cell-cell interactions of MDK signaling pathway in primary and recurrent cSCC, the link size represents the interaction strengthen. **c** The IHC staining of VIM and MDK in representative samples. **d** VIM and TGFB1 in hypodermis (number of positive cell/area), different tissue sites were marked by different color. **e** The IHC staining of MDK in representative samples. **f** The IHC score of MDK in primary, the first and second recurrence of cSCC, different tissue sites were marked by different color. **g** Scatter plot of the score of MDK and VIM, MDK and TGFB1 in tumor samples of cSCC. Different dots represent the type of sample. Wilcoxon signed-rank test, \**p* < 0.05, \*\**p* < 0.01, \*\*\*\**p* < 0.0001.

Next, we focused on TSKs and identified increased interaction signalling in recurrent cSCC using primary cSCC as a control. Among the significantly upregulated pathways, we observed that the MDK pathway was exclusively present in recurrent cSCC (Fig. S9b). Interestingly, primary and recurrent cSCC possessed distinct MDK- associated interaction relationships. In primary cSCC, iCAFs and IL7R**^+^** CAFs produced high levels of MDK and mediated their strong intercellular communications, while in recurrent cSCC, TSKs secreted high levels of MDK, whose receptors were expressed on themselves and other cells, thus regulating the strong cell–cell interactions within TSKs (Fig. 6b, Fig. S9c, Fig. S10). To examine the effect of MDK on fibroblasts, we investigated the expression of MDK and VIM (a general marker of fibroblasts) in normal skin and actinic keratosis (AK) samples. Moreover, we validated the expression of MDK by IHC staining using 5 normal skin samples, 10 samples with AK, and 16 patients with primary tumours and single or multiple recurrences of cSCC, which comprised matched ANS and tumour samples (Materials and methods, Table S1). Compared with normal skin, VIM was upregulated in the hypodermis of AK, indicating the presence of increased fibroblasts in the hypodermis (Fig. 6c-d). MDK was also highly expressed in the hypodermis of AK and displayed a positive correlation pattern with VIM (R = 0.58, *p* = 0.088, Fig. S11a). Therefore, we hypothesized that cSCC has increased fibroblasts in early carcinogenesis, which may be mediated by MDK. Surprisingly, MDK also participated in the recurrence of cSCC, and tumour tissues with the first and second recurrence had higher MDK expression levels than primary cSCC (Figure 6e-f). In addition, MDK was significantly correlated with the expression of VIM and TGFB (Fig. S11b), demonstrating that it may regulate the EMT of cSCC. MDK was demonstrated to be abnormally expressed in human malignancies and participate in diverse cancer development and progression processes(Filippou et al., 2020). This result suggests that MDK may be a double-edged sword in the TME when it is expressed on different cells. When fibroblasts highly express MDK, it may promote the cell replication of fibroblasts and block the proliferation and metastasis of tumour cells through cellular interactions, thus avoiding tumour recurrence. On the other hand, in recurrent tumour tissues, TSKs may acquire proliferative or EMT capacity by expressing MDK, which then mediates tumour recurrence.

Taken together, we found that primary cSCC was similar to a “hot” tumour, which infiltrated with a higher abundance of T cells, whereas recurrent cSCC tended to be a “cold” tumour and harboured EMT characteristics. CD8^+^ T cells secreted high expression of CTLA4 and TIGIT in primary cSCC, while they highly expressed TIM3, CTLA4 and CXCL13 in recurrent cSCC. Recurrent cSCC showed relatively high proportions of SPP1^+^ CD209^high^ and SPP1^+^ CD209^low^ TAMs, which were characterized by marked features of phagocytosis and angiogenesis, respectively. They have less potential for phagocytosis and inflammation, as well as more obvious angiogenic characteristics in recurrent cSCC. We observed that MDK might drive the strong intercellular interactions within iCAFs and IL7R**^+^** CAFs in primary cSCC, while TSKs tend to have strong interactions with themselves by secreting MDK. In addition, in recurrent cSCC, IL7R**^+^**CAFs showed obvious interactions with TSKs by expressing EMT-related markers, which may promote the EMT of tumour cells (Fig. 7a).

**Fig. 7.**
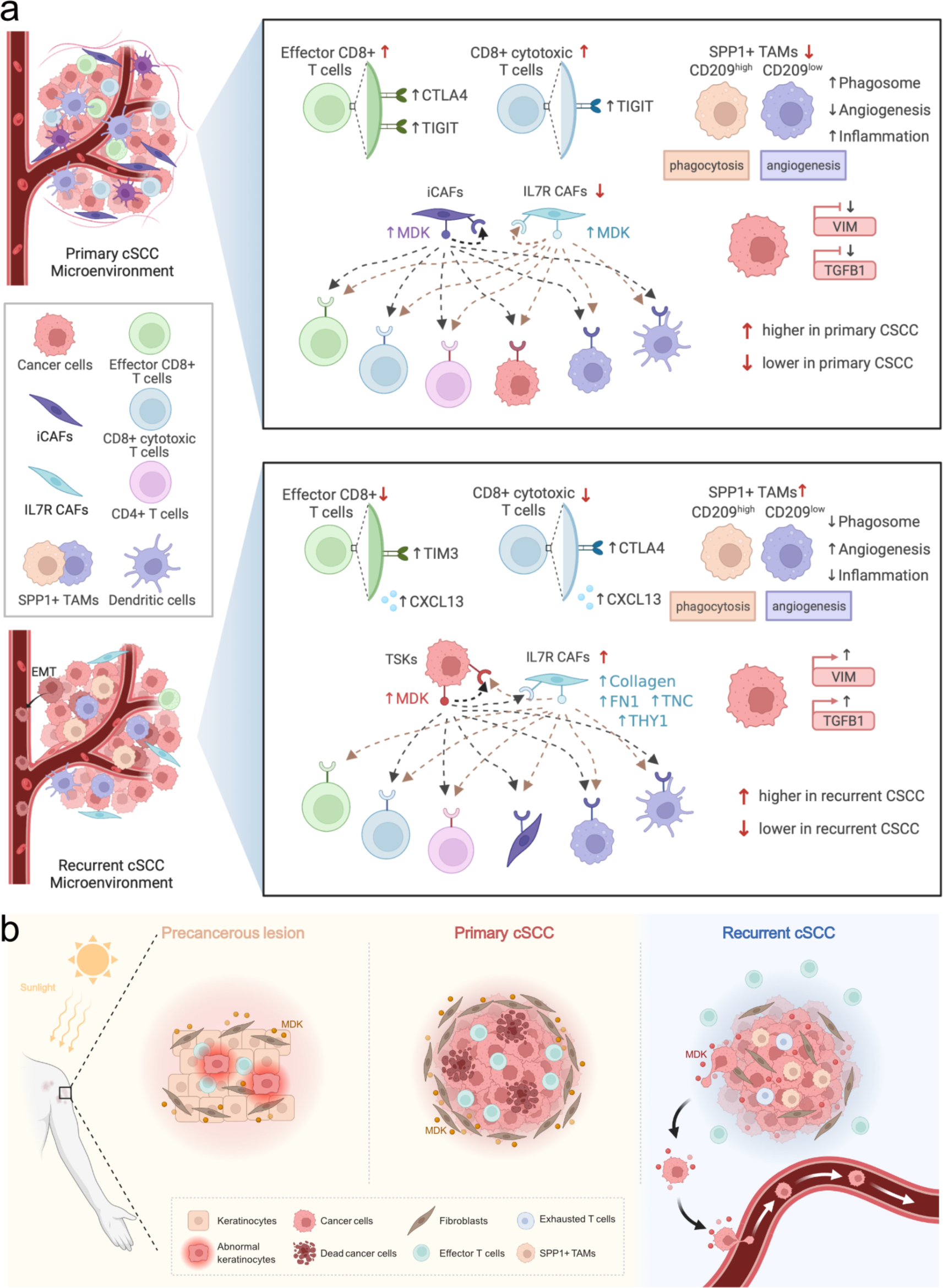
Diagram illustrating the reprogramming of TME in cSCC progression. **a** Top, schematic diagram of the TME and cell-cell interactions in primary cSCC. Bottom, schematic diagram of the TME and cell-cell interactions in recurrent cSCC. **b** The specific transformation pattern of MDK within TME, ranging from a precursor AK, to primary cSCC, and finally recurrent cSCC.

## Discussion

Although cSCC is usually not life-threatening, and most cSCCs can be successfully eradicated by surgical resection, a subset of cSCCs suffer from recurrence, metastasis, and death(Que et al., 2018). The highly complex and heterogeneous microenvironment of a tumour is essential to cancer cell progression and metastasis(Baghban et al., 2020). Here, we performed scRNA-seq on 14,626 single cells of ANS and tumour tissues from 5 patients who were diagnosed with primary or recurrent cSCC. Through integrated analyses of scRNA-seq data, we provided a comprehensive functional and cell–cell interaction landscape of epithelial cells, fibroblasts, myeloid cells and T cells, which are the major components of the TME of cSCC. Our study characterized the heterogenous and functional differences in the TME between primary and recurrent cSCC, and the critical reprogramming of the TME depicted in our study revealed the mechanism of cSCC recurrence, which may provide instructive guidance for clinical prevention and treatment.

In our study, we found that recurrent cSCC was characterized by a low T-cell infiltration proportion and an elevated T-cell exhaustion state. Tumour-infiltrating CD8^+^ T cells have been shown to progress into an exhaustion state within many tumours(Zebley and Youngblood, 2022), such as hepatocellular carcinoma(Ma et al., 2019a), melanoma(Li et al., 2019) and breast cancer(Egelston et al., 2022). High expression of immune checkpoint receptors is associated with the exhaustion state of CD8^+^ T cells, such as PD-1, CTLA-4, TIM-3, and LAG-3(Ma et al., 2019b), and their dysfunctional state has been demonstrated to affect the postoperative survival and recurrence risk of tumour patients(Ma et al., 2019a). In addition, tumours characterized by a lack of effector T cells have been termed “cold tumours” and have been shown to resist immunotherapy(Zhang et al., 2020), indicating that immunotherapy is not suitable for recurrent cSCC. Inflammatory pathways play important roles in regulating the innate and adaptive immune response. TNF (tumour necrosis factor) and NF-κB signalling can promote the activation of effector T cells(Liu et al., 2017; Mehta et al., 2018). IL-17 is a proinflammatory cytokine secreted by T cells that plays critical roles in host defence against bacterial infection(Qian et al., 2010). The NOD-like receptor signalling pathway participates in the regulation of the host innate immune response(Franchi et al., 2009). In particular, T cells in recurrent cSCC harboured decreased activity among the TNF, IL-17, NF-kappa B and NOD-like receptor signalling pathways, which may reflect their low defence and weak killing ability.

A recent study revealed that SPP1^+^ macrophages may prevent the infiltration of lymphocytes, which further reduces the efficacy of PD-L1 treatment(Qi et al., 2022). Moreover, SPP1^+^ macrophages were reported to carry out angiogenesis, which was positively correlated with EMT markers and related to tumour metastasis(Georgoudaki et al., 2016; Cheng et al., 2021; Liu et al., 2022; Qi et al., 2022). Herein, SPP1^+^ TAMs tended to express a global protumour characteristic in recurrent cSCC, including lower phagocytosis and inflammation scores, as well as higher angiogenesis scores, which may be the critical mechanism of cSCC recurrence. However, the two subpopulations of SPP1^+^ macrophages (SPP1^+^ CD209^high^ and SPP1^+^ CD209^low^) possessed different characteristics, showing remarkable features of phagocytosis and angiogenesis, respectively. This indicates the heterogeneous property of SPP1^+^ TAMs, and the proportion of SPP1^+^ CD209^high^ and SPP1^+^ CD209^low^ TAMs may affect the outcomes of cSCC. Patients might have a good prognosis if they contain high proportions of SPP1^+^ CD209^high^ TAMs with strong phagocytosis ability. Surprisingly, we found that recurrent cSCC showed significantly higher expression levels of two EMT-associated genes, VIM and TGFB1, as well as high EMT activity characteristics, which may be related to the proportion of SPP1^+^ TAMs. The high expression of VIM and TGFB1 in recurrent cSCC was further validated using IHC in an independent clinical cohort. The TME is a heterogeneous collection of multiple cells and noncellular tissue components; among them, CAFs act as a double-edged sword with tumour-restraining/promoting roles, which could induce EMT in diverse cancers(Fiori et al., 2019; Goulet et al., 2019; Tyler and Tirosh, 2021). Here, we found that IL7R**^+^** CAFs were enriched in recurrent cSCC and could interact with TSKs by expressing integrins through the COLLAGEN, FN1, TENASCIN and THY1 signalling pathways. Collagen and tenascin C (TNC) can promote EMT in tumours(Shintani et al., 2008; Nagaharu et al., 2011); therefore, the interaction between TSKs and IL7R**^+^** CAFs may mediate the EMT of tumour cells in recurrent cSCC.

MDK is a heparin-binding growth factor that promotes the proliferation and EMT of tumour cells(Filippou et al., 2020). It has been demonstrated to correlate with poor prognosis and promote tumour progression in glioblastoma(Hu et al., 2021). In addition, it could lead to immunotherapy resistance and promote immunosuppression in human cancer (2020). AK is the most common form of precancerous lesion in cSCC, which is related to the cumulative ultraviolet (UV) exposure from sunlight(Ratushny et al., 2012). We measured the abundance of VIM in normal skin and AK, which is a general marker of fibroblasts, and found that it was significantly enriched in the hypodermis of AK. In addition, MDK is also upregulated and tends to have an expression pattern consistent with that of VIM in AK. Researchers isolated fibroblasts from primary cSCC and healthy dermis and observed that fibroblasts derived from cSCC have increased proliferation compared to normal fibroblasts(Commandeur et al., 2011). In our study, the high expression of MDK in fibroblasts led to strong intercellular communication in primary cSCC. This evidence suggests that MDK may drive the massive proliferation of fibroblasts in AK under cumulative exposure to sunlight, which may be a protective mechanism against UV. In addition, when AK progresses into primary cSCC, MDK may further promote fibroblast proliferation, allowing it to regulate ECM remodelling and protect tumour cells from escaping the microenvironment. This hypothesis explains why primary cSCC is unlikely to metastasize, which is based on clinical observation. Astonishingly, in recurrent cSCC, MDK was expressed at low levels in fibroblasts and shifted to a high expression pattern in cSCC, which further promoted cell–cell communication within themselves. In addition, MDK was positively correlated with VIM and TGFB1 in a cSCC cohort, demonstrating that it is associated with EMT. Therefore, MDK potentially drives the proliferation and EMT of tumour cells in recurrent cSCC. Our study proposes a specific transformation pattern of MDK, ranging from a precursor AK to primary cSCC and finally recurrent cSCC, providing a novel potential treatment target in cSCC (Fig. 7b).

## Materials and methods

### Patients and sample collection

Five patients that pathologically diagnosed with cSCC at Xiangya Hospital of Central South University were investigated in this study, including four primary cSCC patients and one recurrent cSCC patient. Fresh tumour and adjacent skin samples from primary and recurrent cSCC were surgically resected from the above-described patients (Table S2). All subjects provided written informed consent, and this study was approved by the institutional ethics committee of Xiangya Hospital of Central South University (2022020109).

### Preparation of single-cell suspensions

The fresh tissues were stored in the MACS Tissue storage solution (Miltenyi Biotec, 130-100-008) on ice after the surgery within 30 mins. Wash the specimens with Hanks Balanced Salt Solution (HBSS) for three times before dissociation. The Human Tumor Dissociation Kit (Miltenyi Biotec, 130-095-929), gentleMACS Dissociator (Miltenyi Biotec, 130-093-235) and gentle MACS C Tubes (Miltenyi Biotec, 130-093-237) were used for obtaining single-cell suspensions, Dead Cell Removal Kit (Miltenyi Biotec, 130-090-101) was used to improve cell viability to meet the requirements of single cell sequencing.

### Single-Cell RNA sequencing and Library preparation

BD Rhapsody system and Singleron platform were used in our study.

#### Sequencing with Singleron platform

Single-cell suspensions (1×10^5^ cells/ml) with PBS (HyClone) were loaded into microfluidic devices using the Singleron Matrix^®^ Single Cell Processing System (Singleron). Subsequently, the scRNA-seq libraries were constructed according to the protocol of the GEXSCOPE^®^ Single Cell RNA Library Kits (Singleron) (Dura et al., 2019). Individual libraries were diluted to 4 nM and pooled for sequencing. At last, pools were sequenced on Illumina HiSeq X with 150 bp paired end reads.

#### Sequencing with BD Rhapsody system

Cells from each patient were labeled with sample tags from the Human Immune Single-Cell Multiplexing Kit (BD Biosciences) according to the manufacturer’s instruction. The cell capture beads were retrieved for reverse transcription as per the manufacturer’s protocol (BD Biosciences, Single-Cell Capture and cDNA Synthesis). Single-cell capture and cDNA library preparation were performed using the BD Rhapsody Express Single-Cell Analysis System (BD Biosciences), following the manufacturer’s instructions. The libraries were loaded on an S1 flow cell (2 ×100 cycle) and paired-end sequenced at >200,000 reads per cell depth on a Novaseq 6000 Sequencer (Illumina) at the Kinghorn Centre for Cellular Genomics, Garvan Institute, Sydney, Australia. PhiX (20%) was added to the sequencing run to compensate for the low complexity library(Mayer et al., 2021).

### Raw data processing of single-cell RNA sequencing data

For data generated with Singleron platform, reads were mapped to the human genome (GRCh38) using scopetools (https://github.com/SingleronBio/SCOPE-tools). First, cell barcode and unique molecular identifier (UMI) were extracted after filtering read one according to the sequence information, the corrected barcode and original UMI sequence were added to the ID of read two. After that, read two were trimmed by cutadapt and then align to the reference genome using STAR(Dobin et al., 2013; Kechin et al., 2017). Furthermore, featureCounts was applied to target reads to the genomic position of genes (ensemble version 99)(Liao et al., 2014). Finally, reads with the same cell barcode, UMI and gene were grouped together to count the number of UMIs per gene per cell.

For data generated with BD platform, the FASTQ files were processed following the BD Rhapsody Analysis pipeline (BD Biosciences), which was implemented in the CWL-runner. Briefly, read pairs with low quality were removed, the quality-filtered R1 reads were analysed to identify cell label and UMI sequences. Next, the pipeline uses STAR to map the filtered R2 reads to the transcriptome. Reads with the same cell label, the same UMI sequence and the same gene were collapsed into a single raw molecule. The obtained counts were adjusted by BD Biosciences-developed error correction algorithm—recursive substitution error correction (RSEC) to correct sequencing and PCR errors. Barcoded oligo-conjugated antibodies (single-cell multiplexing kit; BD Biosciences) were used to infer the origin of sample by the BD Rhapsody Analysis pipeline.

### Quality control, batch correction, and major cell type annotation of single-cell RNA sequencing data

The UMI count data of Singleron and BD platform were imported into Seurat (V4.1.0). The following initial cell filtering steps were performed: 1) cells with more than 15% mitochondrial counts were removed; 2) cells expressing less than 200 genes were removed; 3) cells expressing more than 25,000 UMIs were removed. Putative doublets were removed for each sample using the Scrublet tool with the default parameters(Butler et al., 2018). Single-cell gene expression data was normalized using the “LogNormalize” method with a scale factor 10,000. Next, the top 2,000 most variable genes were identified and a linear scaling method was applied to standardize the range of expression values for each gene. Principal component analysis (PCA) was performed to reduce dimensionality by “RunPCA” function. The top 50 principal components (PCs) were used for Uniform Manifold Approximation and Projection for dimension reduction (UMAP). To integrate cells from different samples and platforms for unsupervised clustering, we used harmony and set sample and platform as two technical covariates for batch correction(Korsunsky et al., 2019). Cell clusters were identified using the “FindClusters” function, with the resolution of 0.04. The most significant differentially expressed genes (DEGs) in each cluster were identified using the “FindAllMarker” function, using the Wilcoxon’s test. We further identified major cell types according to the gene expression of well-known markers: Epithelial cells: KRT14, KRT5; T cells: CD2, CD3D; Fibroblasts: COL1A1, DCN; Myeloid cells: LYZ, HLA-DRA; Endothelial cells: RAMP2, VWF; Mast cells: CPA3, KIT; B cells: MZB1, CD79A.

### Sub-clustering of major cell types

For the epithelial cells, T cells, fibroblasts and myeloid cells, we extracted cells from the integrated dataset for sub-clustering. Gene re-scaling, dimensionality reduction, batch correction and cell clustering were performed as described above.

For epithelial cells, we examined the following well-known markers and divided them into 4 cell types: Cycling cells: MKI67, TOP2A; TSKs: PTHLH, MMP10; Basal cells: COL17A1; Differentiating cells: KRT1, KRT16. The Cycling cells were further clustered into cycling basal cells and cycling TSKs based on the above markers. Specifically, we removed cell clusters that highly expressed the cell markers of Pilosebaceous/Eccrine (SAA1, LHX2), fibroblasts (COL1A1), T cells (CD2, CD3D), Mast cells (TPSAB1, TPSB2) and Myeloid cells (LYZ), which were regarded as doublets (Supplementary Figure 2). Few cells (2 cells) from the ANS were clustered into TSK, which may be caused by sample contamination, we thus removed these cells in the downstream analysis.

For T cells, cell subpopulations were determined according to the following gene markers: Effector CD4 T cells: CD4; Naïve CD4^+^ T cells: CD4, CCR7, SELL; Effector CD8 T cells: CD8A, GZMA; CD8^+^ cytotoxic cells: CD8, IFNG, GZMA; Tregs: CD4, IL2RA, FOXP3, CTLA4.

The myeloid cells were sub-clustered based on the gene expression of the following markers: Monocytes: VEGFA, VCAN, FCN1; SPP1^+^ CD209^high^ M2 macrophages: SPP1, CD209, CD163, MRC1, CCL18; SPP1^+^ M2c macrophages: SPP1, CD209, CD163, MRC1, CCL18; M2d macrophages: CD163, MRC1, CXCL10; Cycling M2 macrophages: CD163, MRC1, TOP2A, MKI67; CD14^+^ DCs: CD1A, CD14, CD1C; CLEC9A^+^ DCs: CLEC9A, CD1C; CD1a^+^ CD1c^+^ DCs: CLEC10A, CD1A, CD1C; CXCL9-11^+^ MDSCs: CXCL9, CXCL10, CXCL11, IL1B, S100A8, S100A9; CXCL1-3^+^ MDSCs: CXCL1, CXCL2, CXCL3, IL1B, S100A8, S100A9.

For fibroblasts, cell subpopulations were determined according to the gene expression of the following markers: mCAFs: RGS5, DCN, COL6A2, COL1A1, COL1A2; iCAFs: RGS5, DCN, COL6A2, COL1A1, COL1A2; IL7R**^+^** CAFs: IL7R, IL1B, IL6, CXCL1, CXCL3, CXCL5, CXCL6, CXCL8, CXCL13, CXCL14, DCN, COL6A2, COL1A1, COL1A2.

### Correlation analysis between different cell types

To explore the correlation between different cell types, we first calculated the mean gene expression level of cells that belong to the same cell type, and merged them to computed spearman correlation coefficient. Pheatmap package (V1.0.12) was used to visualized the correlation coefficient between different cell types.

### Identification of differentially expressed genes

The “FindMarkers” function in Seurat package was used to detect differentially expressed genes, we define genes with p_val_adj < 0.05, avg_log2FC > 1 as up- regulated genes and genes with p_val_adj < 0.05, avg_log2FC < -1 as down-regulated genes.

### Cell-cell communication analysis

The CellChat (V1.1.3, https://github.com/sqjin/CellChat) algorithm was used to infer cell-cell interactions within TME and identify differential interactions between primary and recurrent samples(Jin et al., 2021). In brief, we followed the official workflow and imported gene expression data of cSCC into CellChat using “createCellChat” function. We mainly applied “identifyOverExpressedGenes”, “identifyOverExpressedInteractions”, “projectData” functions to detect significant cell-cell interactions among the investigated cells. The “compareInteractions”, “RankNet” functions were used to perform interaction comparison between primary and recurrent samples. The “netAnalysis_signalingRole_scatter” function was used to calculate the incoming and outgoing interaction strengthen of cells among datasets. All cell interaction visualizations were plotted using the CellChat package.

### Gene set variation analysis

#### Hallmarks of cancer activity analysis

We collected GO terms mapping to the hallmarks of cancer(Plaisier et al., 2012), which were used to evaluate the tumour property of TSKs. We applied GSVA method to calculate the hallmark score of individual cells, as implemented in the GSVA R package (V1.40.1)(Hanzelmann et al., 2013).

#### Pathway and gene signature score analysis

We obtained genes from four pathways derived from Kyoto Encyclopedia of Genes and Genomes (KEGG) to evaluated the inflammatory score: TNF signaling pathway (hsa04668), NF-kappa B signaling pathway (hsa04064), IL-17 signaling pathway (hsa04657) and NOD-like receptor signaling pathway (hsa04621). Besides, genes in cMAP signaling pathway (hsa04024) were used to calculate cMAP activity score. We also evaluated the phagocytosis score of myeloid cells by collecting its associated genes from KEGG (hsa04666). The EMT signature related genes were downloaded from MSigDB(Liberzon et al., 2011), which were used to evaluated the EMT score of TSKs. The score of above-mentioned pathways or signature was also calculated using GSVA R package (V1.40.1). To compare the differences of scores between different sample or cell types, we used the Wilcoxon signed-rank test that implemented in the ggpubr package (V0.4.0) to perform significance tests.

### Gene set enrichment analysis

We performed KEGG enrichment analysis using the differentially expressed genes by clusterProfiler (V4.1.4) package(Yu et al., 2012; Wu et al., 2021b). Pathways with *Q* value < 0.05 were regarded as significant enriched results. Besides, we also did gene set enrichment analysis (GSEA) of KEGG pathways using clusterProfiler package, a cutoff *Q* value < 0.05 was applied to select the most significantly enriched pathways.

### Definition of exhaustion score for CD8^+^ T cells

To evaluate the exhaustion status of CD8^+^ T cells in our study, we used a group of exhaustion-related genes to define the exhaustion score(Zhang et al., 2021). Specifically, the exhaustion score was defined as the average expression of CXCL13, HAVCR2, PDCD1, TIGIT, LAG3, CTLA4, LAYN, RBPJ, VCAM1, TOX and MYO7A.

Wilcoxon signed-rank test was used to perform significance tests, which is implemented in the ggpubr package (V0.4.0).

### Construction of single-cell trajectories

#### RNA velocity estimation

The bam files generated by Singleron and BD Rhapsody Analysis pipeline were imported into the Velocyto pipeline(La Manno et al., 2018) to generate loom files, which recorded the count matrices for spliced and unspliced reads. The resulting loom files were fed into the scVelo package (V0.2.4)(Bergen et al., 2020) to computed steady-state gene-specific velocities, the final cell transition status was visualized in our original UMAP embedding.

#### CytoTRACE analysis

We performed CytoTRACE(Gulati et al., 2020) analysis with default parameters following the official guidance, which could predict cell differentiation states from scRNA-seq data. The CytoTRACE score was used to verify the trajectory analysis from scVelo package.

### IHC staining in human cSCC samples

For immunohistochemical staining, the samples collected from 16 patients with cSCC (Supplementary Table 2) were sectioned for 4 um. Tissue sections heated (65°C) in an oven for 2 hours, followed by 40 minutes in deparaffinization and rehydrated with graded alcohol concentrations using standard procedures. Immersed in the already boiling 10nM citric acid (pH 6.0), the deparaffinized sections heated on low heat for 20 minutes in a microwave oven. For blocking endogenous peroxidase activity, the deparaffinized sections was incubated with endogenous peroxidase blocking solution for 10 minutes, and then incubated with normal goat serum for blocking for 1 hour to block nonspecific immunoglobulin binding. Then, the slides were incubated at 4°C overnight with a primary rabbit monoclonal antibody against MDK (1:100, ab52637, Abcam), vimentin (1:200, TU253239, abmart), TGF beta 1 (1:200, PU159710, abmart).

The next day, the slides were incubated with IHC enhancer for 20 minutes, followed by incubation with the corresponding secondary antibody conjugated with horseradish peroxidase at 37℃ for 1 hour. Then, DAB (3,3-diaminodbenzidine) substrate was added, and then counterstained with hematoxylin for 5 min. Finally, the sections were dehydrated, cleared and mounted in aqueous mounting medium for microscopic evaluation.

### Semi-quantitative analysis of IHC staining

To achieve high quality results, two independent pathologists experienced in evaluating IHC participated in reviewing samples, who were blinded to the clinical outcome of these patients. We assessed the percentage of positively stained immuno-reactive cells and the staining intensity to semi-quantitatively determine the expression of MDK, VIM and TGFB1. The percentage of immuno-reactive cells was rated as follows: 0 points, <10%; 1 point, 10–50%; 2 points, >50%. The staining intensity was rated as follows: 0 (no staining or weak staining = light yellow), 1 (moderate staining = yellow brown) and 2 (strong staining = brown). The overall score for MDK/VIM/TGFB1 expression was the sum of points determined for the percentage of positively stained immuno-reactive cells and the expression, and an overall score ranging from 0 to 4 was assigned. For the statistical analysis, the patients were divided into a low expression group (an overall score between 0 and 2) and a high expression group (an overall score between 3 and 4)(Yin et al., 2011; Yin et al., 2016; Yin et al., 2020). The final score is the combination of independent scores assigned by the two pathologists, which was reported in this study. Any differences in the scores were resolved by discussion between the two pathologists.

### Data availability

The IHC results generated during this study are provided as supplementary tables. single-cell transcriptome and additional datasets are available upon request.

## Acknowledgements

This work was supported by grants from General Program, The National Natural Science Foundation of China (No. 81874138 and 82073020), Hunan Natural Science Foundation for Distinguished Young Scholars (No. 2021JJ10073), The Youth Science Foundation of Xiangya Hospital (No. 2021Q10).

## Contributions

M.Y. and X.C. conceived and supervised this project. X. L. S. Z. analyzed the scRNA-seq data. S. Z. collected clinical samples and supervised the experiment. H. B. prepared the single-cell suspensions and did IHC staining. X. L., M.Y., X.C., S. Z. and H. B. wrote the manuscript. L. L., S. P., L. D. organized figures and supervised the experiment. W. S., L. H., X. Z. and M. C. edited the manuscript.

## Corresponding authors

Correspondence to Mingzhu Yin, Xiang Chen.

## Ethics declarations Competing interests

The authors declare no competing interests.

---

**Fig. S1.**
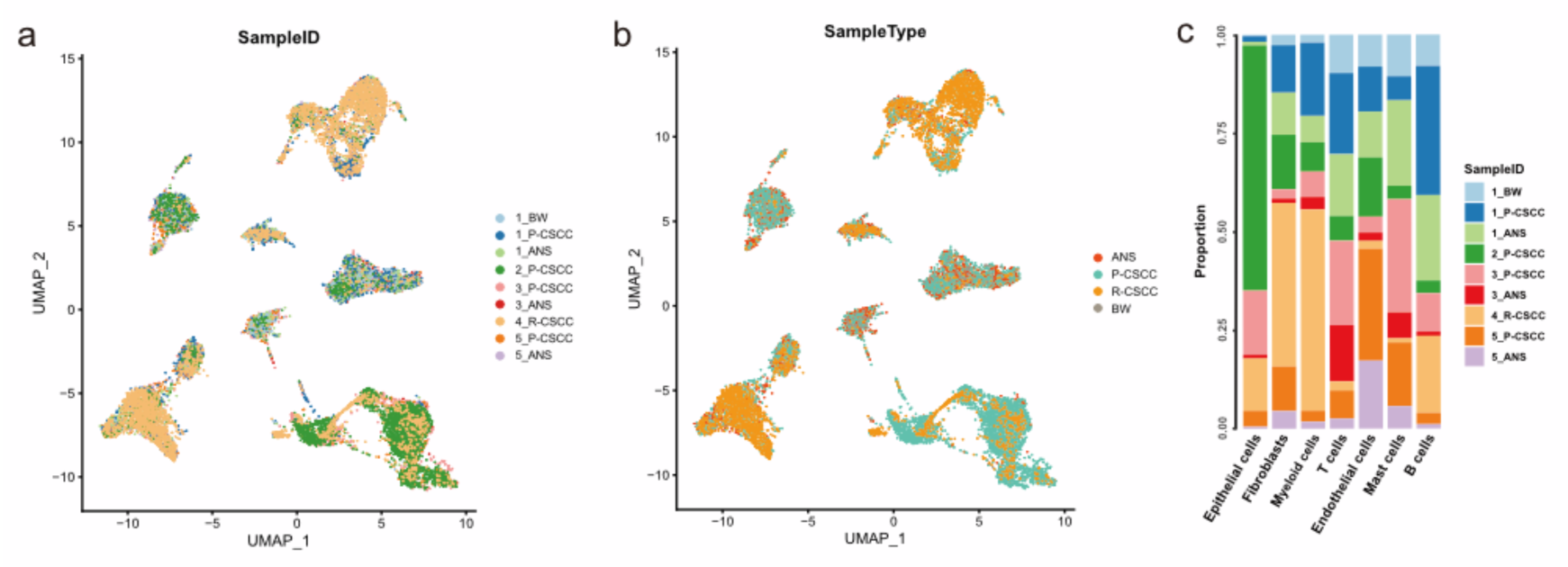
Visualization of single cells profiled in our study. **a** UMAP plot colored by samples. **b** UMAP plot colored by sample types. **c** Sample fractions relative to the total cell count per cell type.

**Fig. S2.**
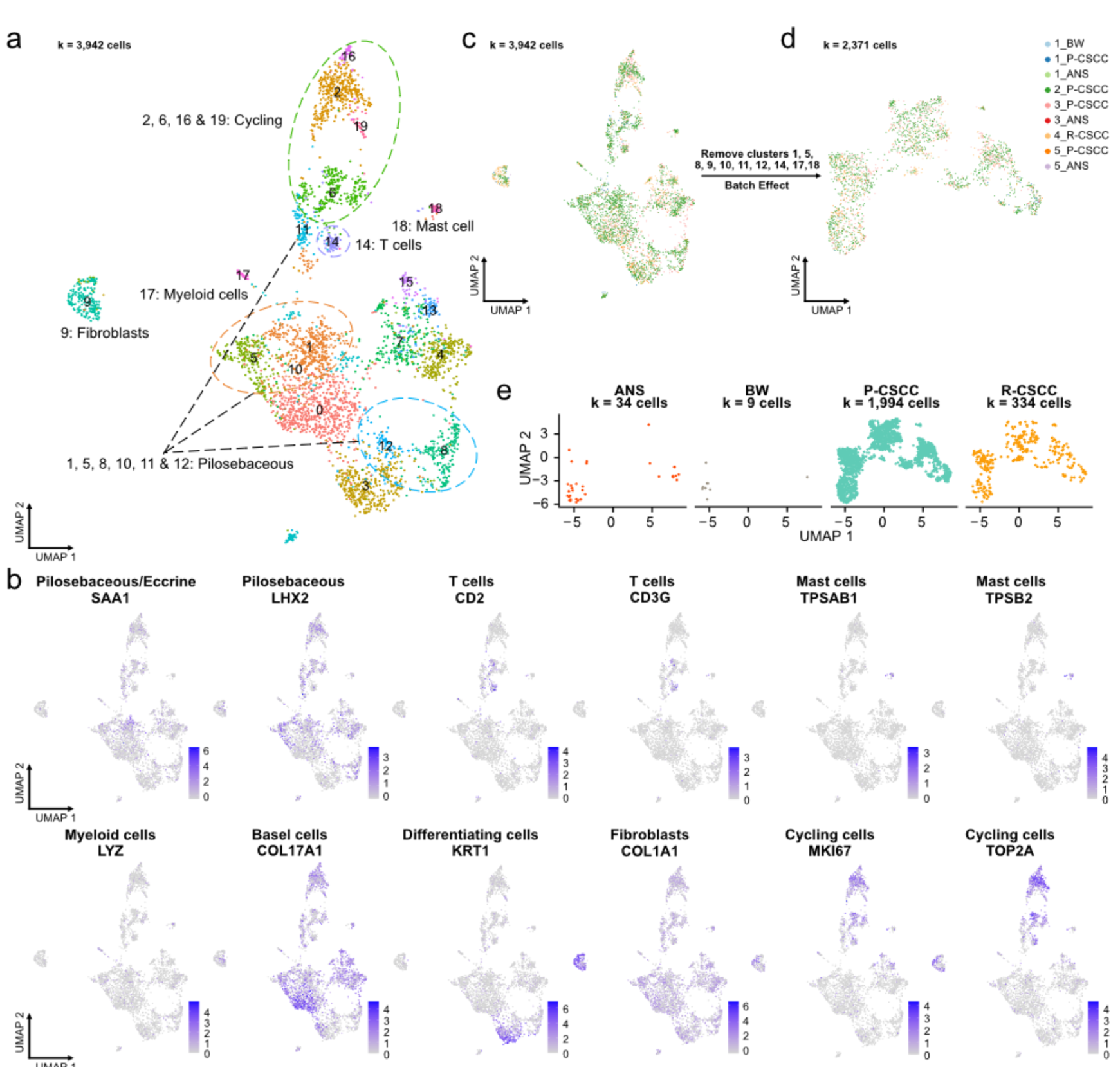
Epithelial cells clustering and annotation. **a** Initial classification results of epithelial cells. **b** The expression of specific cell markers among UMAP. **c** UMAP plot colored by samples before removing doublets. **d** UMAP plot colored by samples after removing doublets, re-scaling, and clustering. **e** UMAP plot colored by sample types after removing doublets, re-scaling, and clustering.

**Fig. S3.**
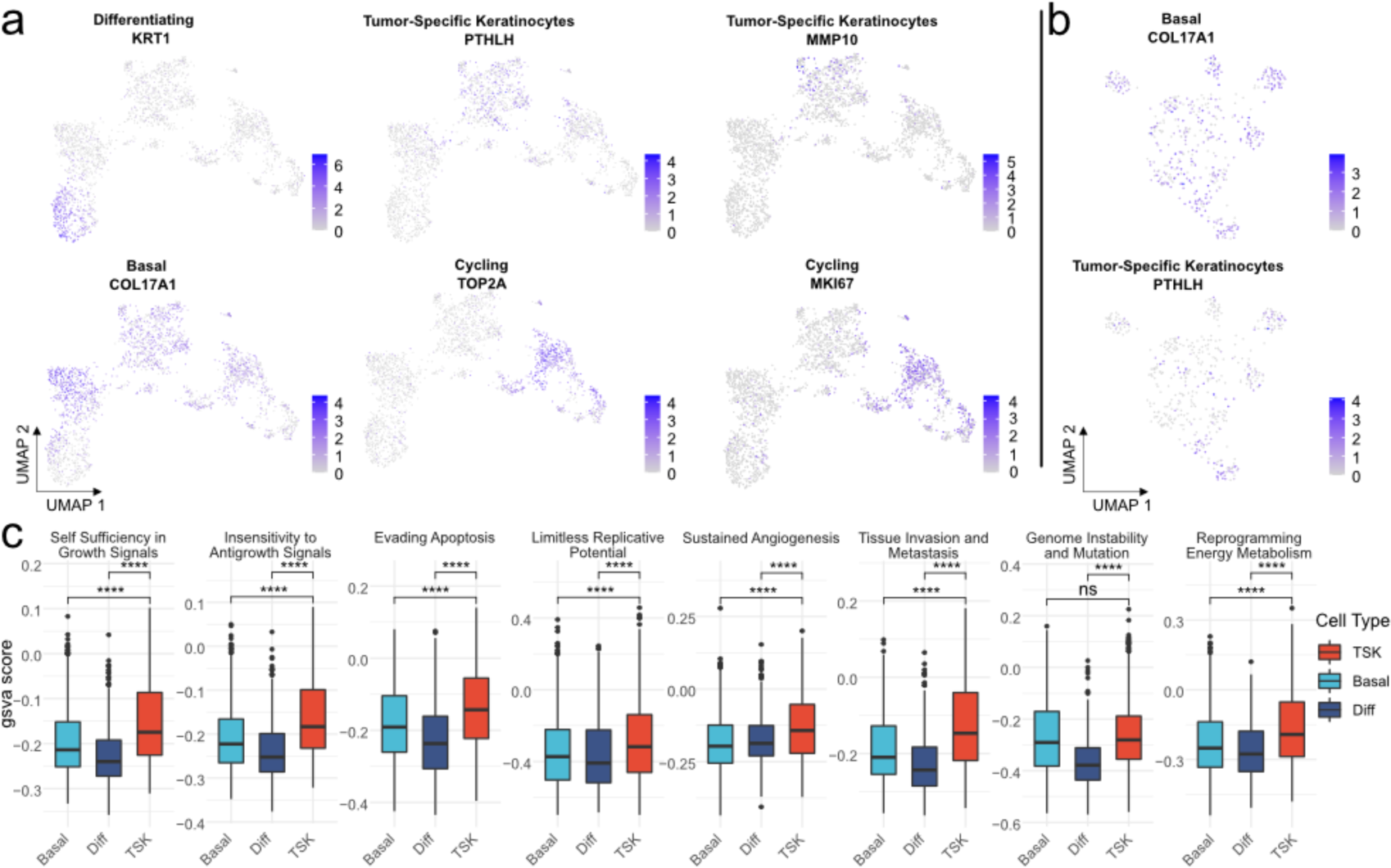
Functional characterization of epithelial cells in cSCC. **a-b** UMAP plot shown the expression level of epithelial cell associated markers. **c** The GSVA scores of “hallmarks of cancer” among different sample types. Wilcoxon signed-rank test, \*\*\*\**p* < 0.0001.

**Fig. S4.**
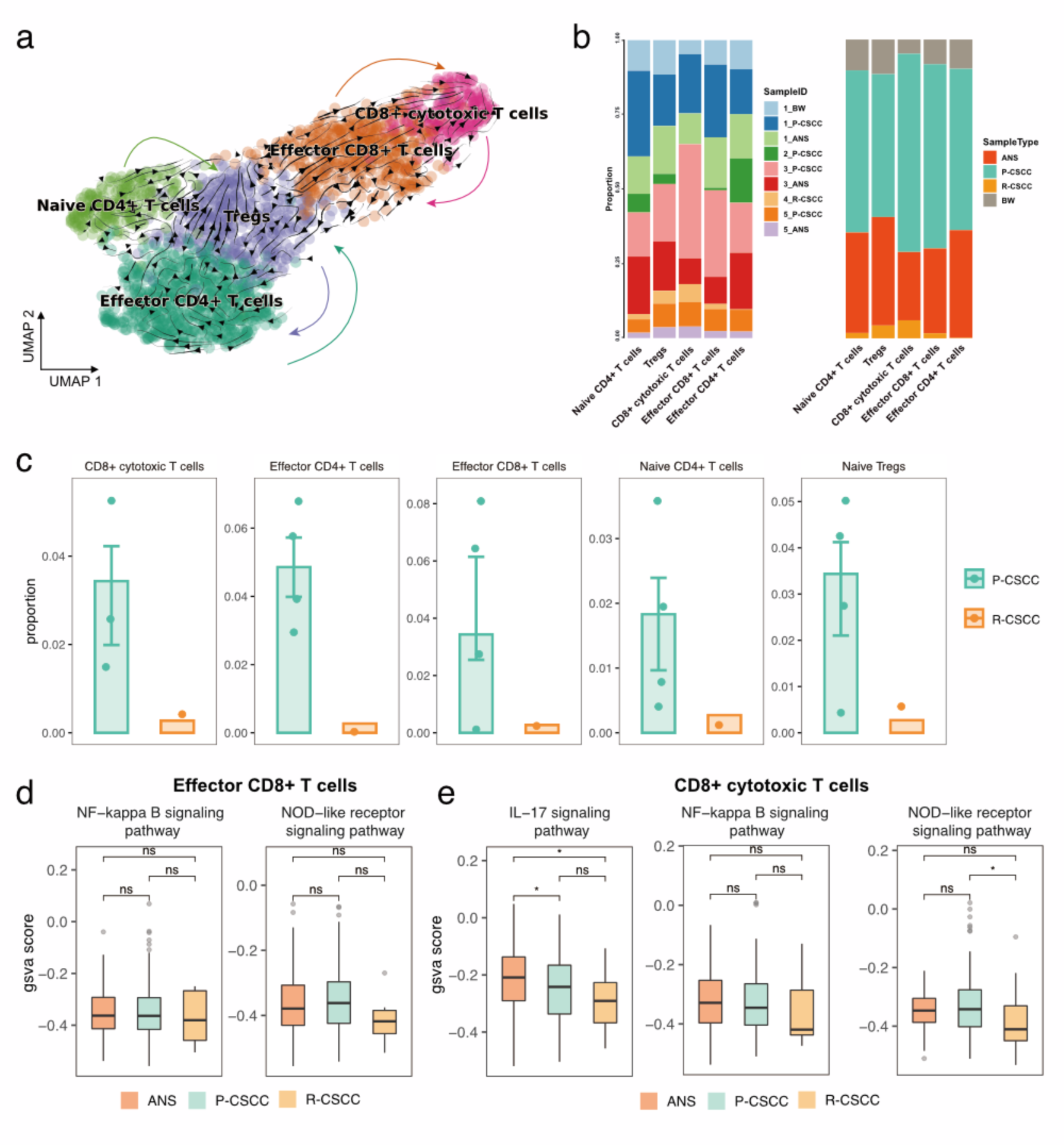
Single-cell transcriptomic analysis of T cells in cSCC. **a** Cell transition potential determined by RNA velocity analysis. **b** The proportion of T cells relative to sample ID and sample type. **c** Each point represents the proportion of T cells among samples, Y-axis represents the average proportion of T cells in each group, Error bars represent ± S.E.M. **d** The GSVA score of inflammatory pathways in effector CD8+ T cells among different sample types. **e** The GSVA score of inflammatory pathways in CD8+ cytotoxic T cells among different sample types. Wilcoxon signed-rank test, \**p* < 0.05.

**Fig. S5.**
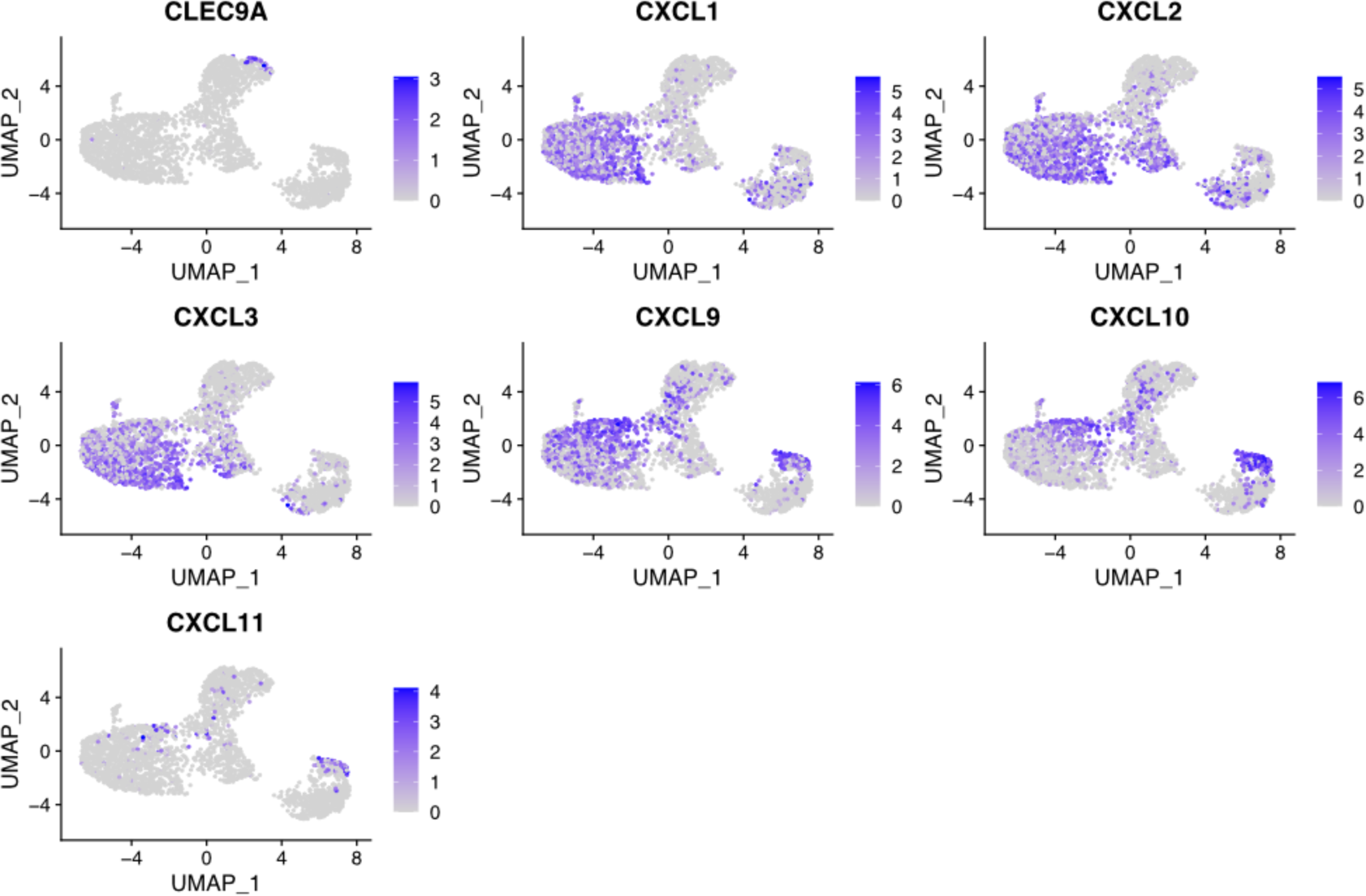
The expression level of gene markers that related with DCs and MDSCs.

**Fig. S6.**
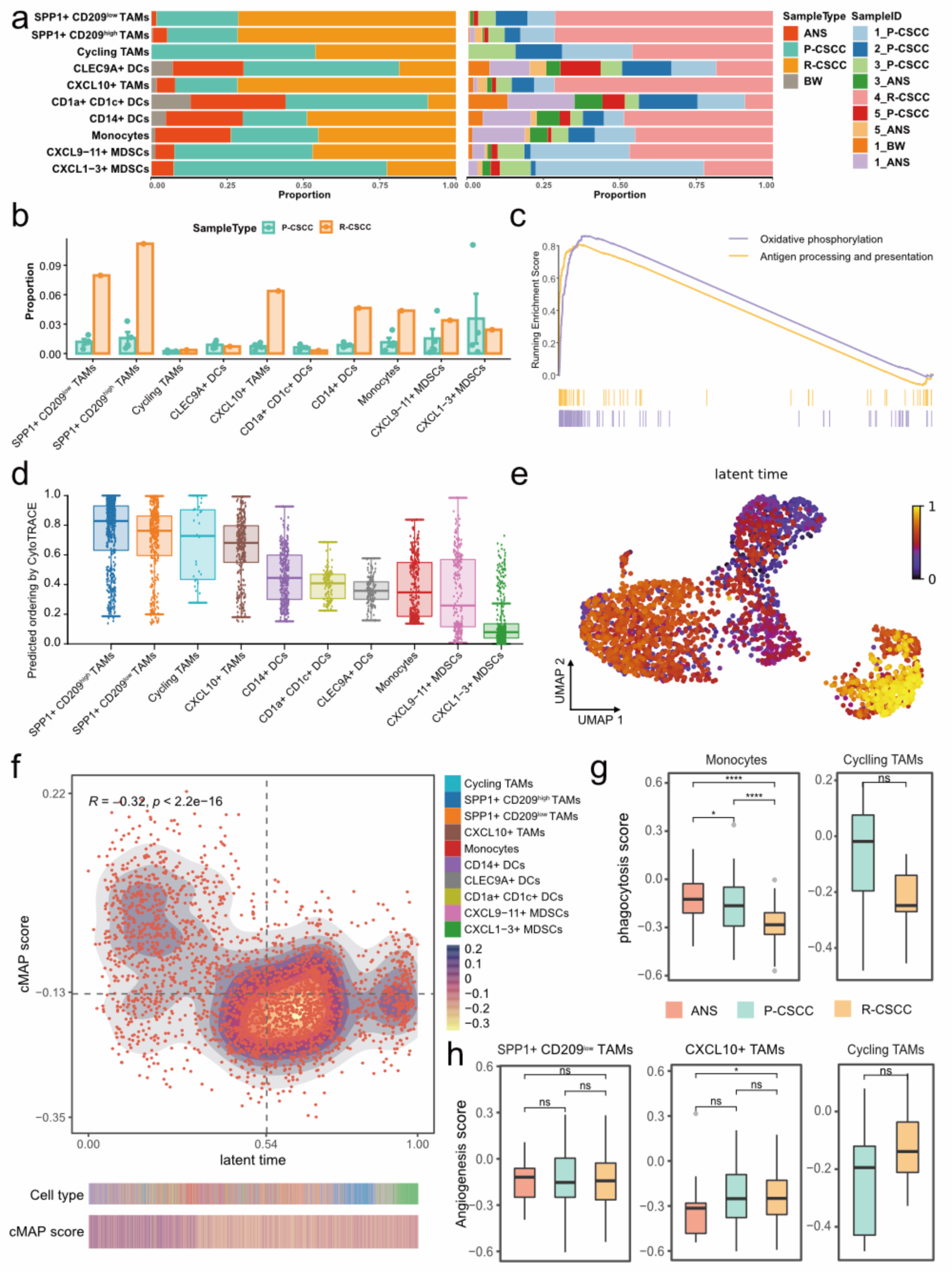
Components and phenotypes of myeloid cells in cSCC. **a** The proportion of myeloid cells relative to sample ID and sample type. **b** Each point represents the proportion of myeloid cells among samples, Y-axis represents the average proportion of myeloid cells in each group, Error bars represent ± S.E.M. **c** Significant enriched pathways in CXCL9-11^+^ MDSCs versus CXCL1-3^+^ MDSCs. **d** Distribution of CytoTRACE score in each cell type, ranking by the median value. **e** UMAP plot showing the latent time estimated by scVelo tool. **f** The top scatter plot showing the relationship between latent time and cMAP pathway score, the bottom heatmap plot showing the cMAP score relative to the latent time. **g** The GSVA score of phagocytosis in monocytes and Cycling TAMs among different sample types. **h** The GSVA score of angiogenesis in SPP1^+^ CD209^low^ TAMs, CXCL10^+^ TAMs and Cycling TAMs among different sample types. Wilcoxon signed-rank test, \**p* < 0.05, \*\*\*\**p* < 0.0001.

**Fig. S7.**
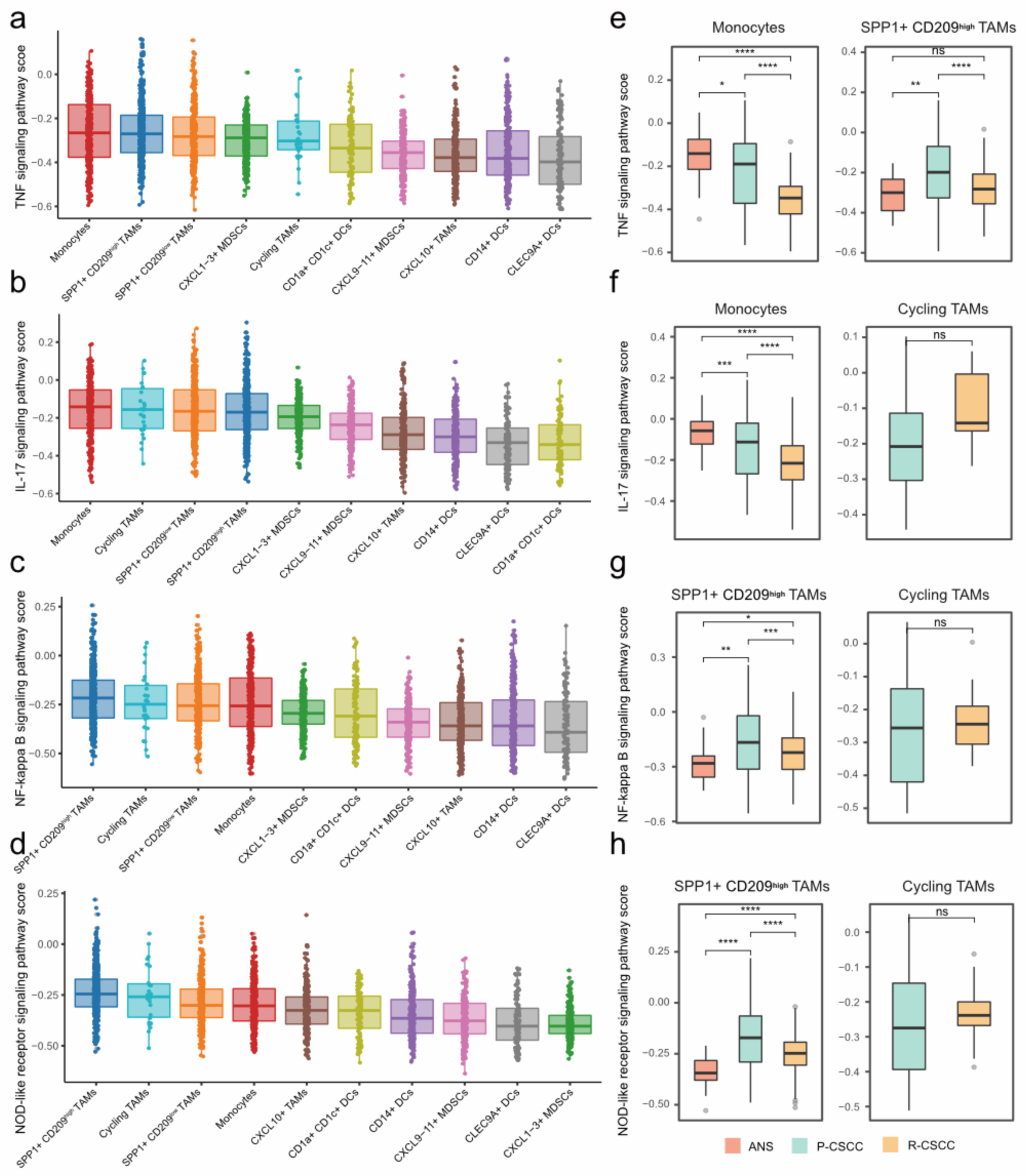
The inflammatory character of myeloid cells in cSCC. **a** Distribution of TNF signaling pathway score in each cell type, ranking by the median value. **b** Distribution of IL-17 signaling pathway score in each cell type, ranking by the median value. **C** Distribution of NF-kappa B signaling pathway score in each cell type, ranking by the median value. **d** Distribution of NOD-like receptor signaling pathway score in each cell type, ranking by the median value. **e** The GSVA score of TNF signaling pathway in monocytes and SPP1^+^ CD209^high^ TAMs among different sample types. **f** The GSVA score of IL-17 signaling pathway in monocytes and Cycling TAMs among different sample types. **g** The GSVA score of NF-kappa B signaling pathway in SPP1^+^ CD209^high^ TAMs and Cycling TAMs among different sample types. **h** The GSVA score of NOD-like receptor signaling pathway in SPP1^+^ CD209^high^ TAMs and Cycling TAMs among different sample types. Wilcoxon signed-rank test, \**p* < 0.05, \*\**p* < 0.01, \*\*\**p* < 0.001, \*\*\*\**p* < 0.0001.

**Fig. S8.**
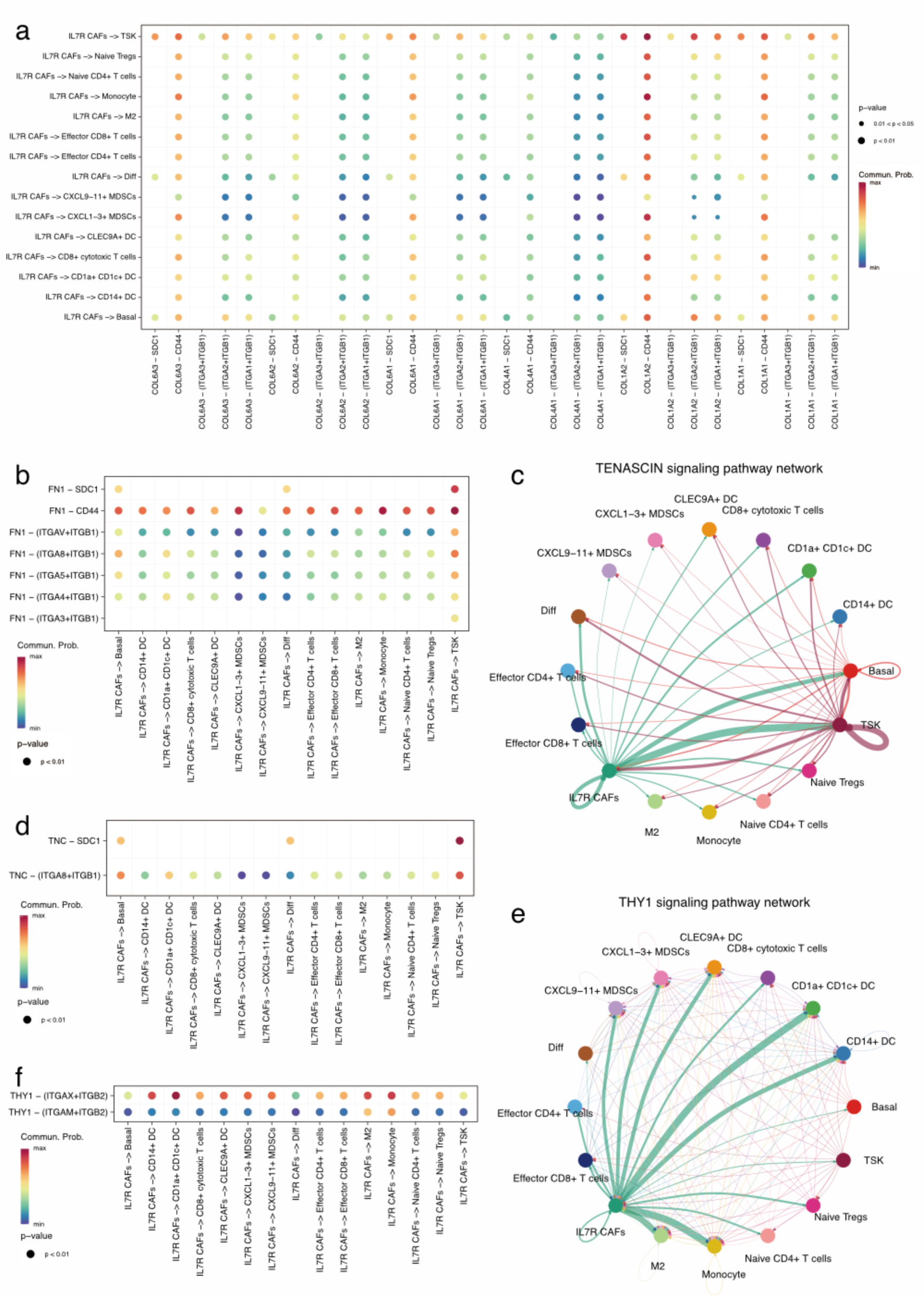
Cell-cell interactions between IL7R^+^ CAFs and other cells. **(a, b, d, f)** Summary of ligand–receptor interactions of COLLAGEN signaling pathway, FN1 signaling pathway, TENASCIN signaling pathway and THY1 signaling pathway. P– values are represented by the size of each circle. The color gradient indicates the communication probability of interaction. **(c, e)** Cell-cell interactions in TENASCIN signaling pathway and THY1 signaling pathway.

**Fig. S9.**
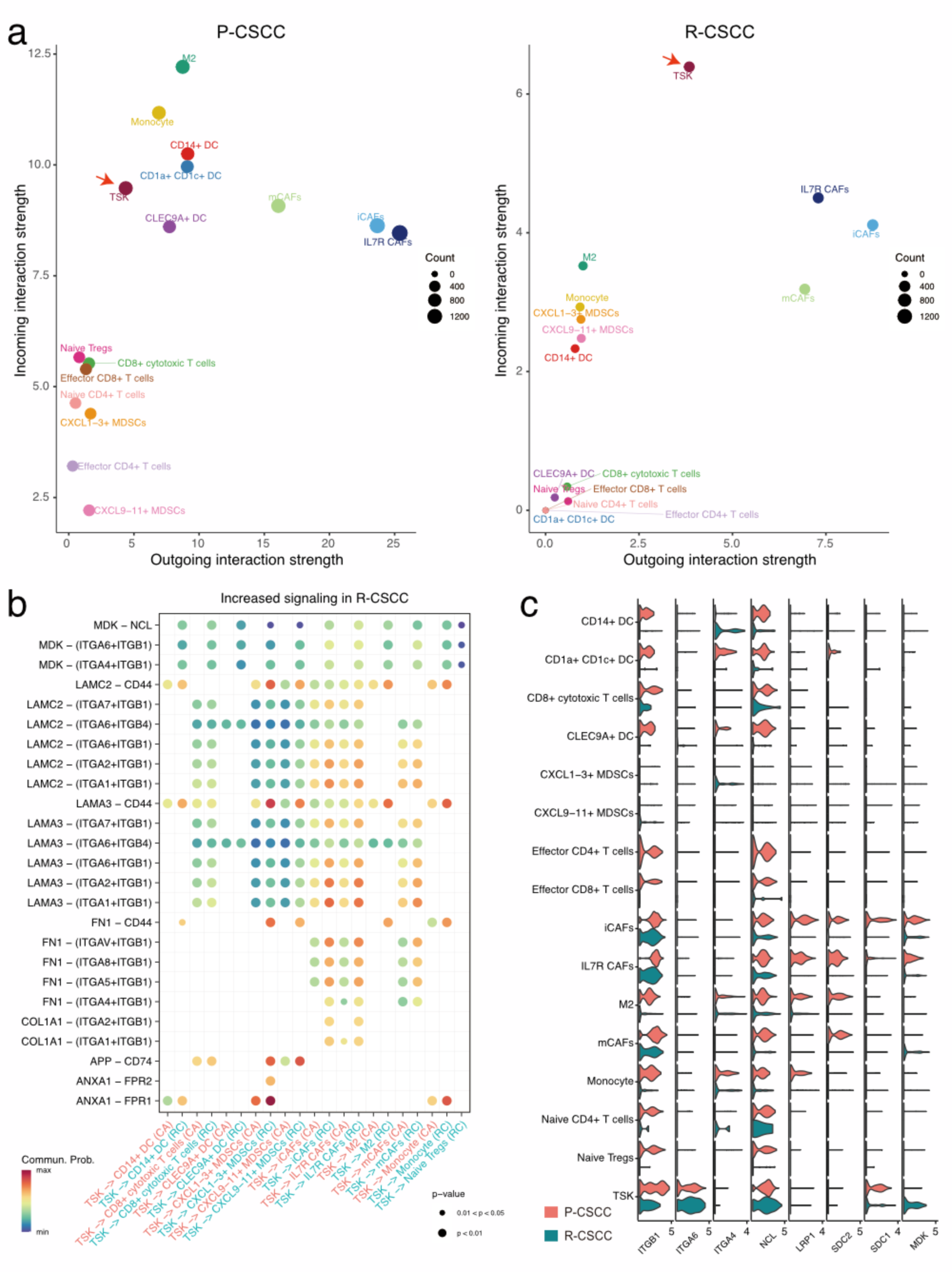
Cell-cell interactions of the primary and recurrent cSCC. **a** The incoming and outgoing interaction strength of different cells in primary and recurrent cSCC. **b** Summary of ligand–receptor interactions of increased signaling pathway in recurrent cSCC. P–values are represented by the size of each circle. The color gradient indicates the communication probability of interaction. **c** The expression of ligands and receptors among different cells, colors represent different sample types.

**Fig. S10.**
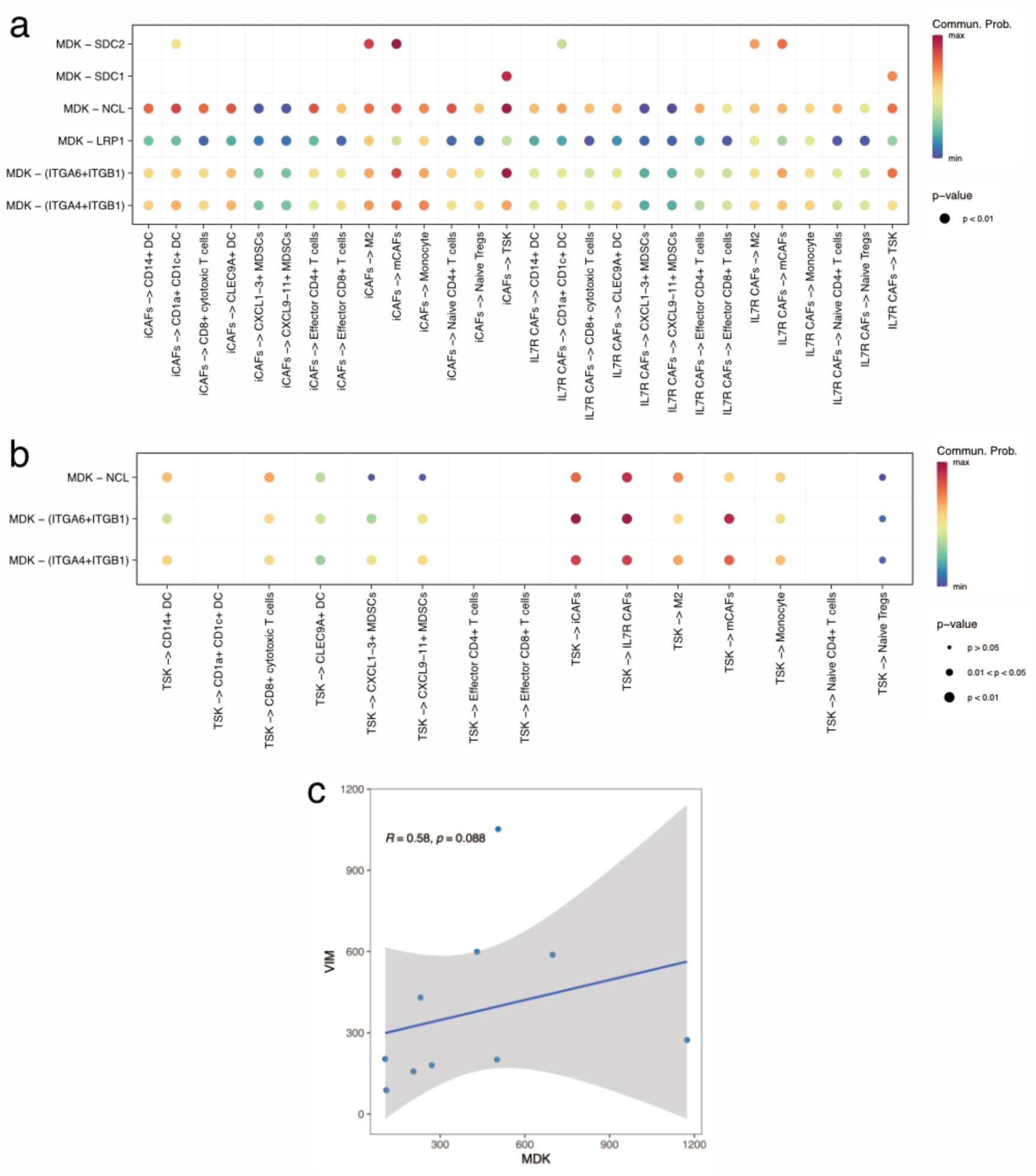
The specific ligand–receptor interactions of MDK signaling pathway within TME and correlation analysis in clinical samples. **a** Summary of ligand– receptor interactions of MDK signaling pathway in primary cSCC. **b** Summary of ligand–receptor interactions of MDK signaling pathway in recurrent cSCC. P–values are represented by the size of each circle. The color gradient indicates the communication probability of interaction. c Scatter plot of the score of MDK and VIM in AK samples.

**Table S1.**
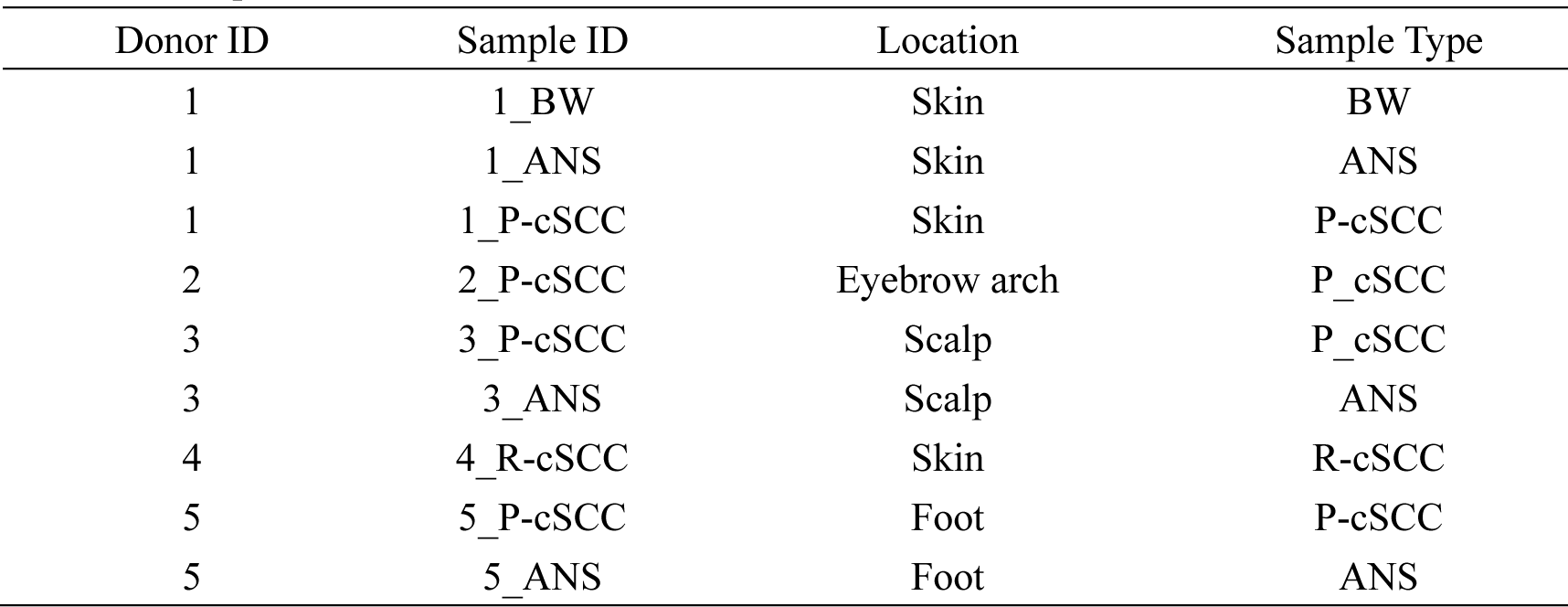
Sample information.

**Table S2.** IHC score of VIM, TGFB1 and MDK.

| Patient ID | Type | VIM (ANS) | VIM (cSCC) | TGFB1 (ANS) | TGFB1 (cSCC) | MDK (ANS) | MDK (cSCC) |
| --- | --- | --- | --- | --- | --- | --- | --- |
| 1 | Primary | 0 | 0 | 0 | 0 | 0 | 0 |
| 1 | Recurrence<sup>1st</sup> | 0 | 2 | 0 | 1 | 1 | 2 |
| 1 | Recurrence<sup>2nd</sup> | 0 | 3 | 0 | 3 | 0 | 4 |
| 2 | Primary | 0 | 1 | 0 | 0 | 0 | 0 |
| 2 | Recurrence<sup>1st</sup> | 0 | 3 | 0 | 4 | 0 | 2 |
| 2 | Recurrence<sup>2nd</sup> | 0 | 3 | 0 | 3 | 1 | 3 |
| 3 | Primary | 0 | 0 | 0 | 0 | 0 | 1 |
| 3 | Recurrence<sup>1st</sup> | 0 | 2 | 0 | 2 | 0 | 3 |
| 3 | Recurrence<sup>2nd</sup> | 0 | 2 | 0 | 3 | 0 | 3 |
| 4 | Primary | 0 | 0 | 0 | 0 | 1 | 2 |
| 4 | Recurrence<sup>1st</sup> | 0 | 1 | 0 | 1 | 0 | 4 |
| 4 | Recurrence<sup>2nd</sup> | 0 | 2 | 0 | 3 | 0 | 3 |
| 5 | Primary | 0 | 0 | 0 | 0 | 0 | 1 |
| 5 | Recurrence<sup>1st</sup> | 0 | 3 | 0 | 3 | 0 | 3 |
| 6 | Primary | 0 | 1 | 0 | 1 | 0 | 0 |
| 6 | Recurrence<sup>1st</sup> | 0 | 3 | 0 | 3 | 0 | 2 |
| 7 | Primary | 0 | 0 | 0 | 1 | 1 | 2 |
| 7 | Recurrence<sup>1st</sup> | 0 | 2 | 0 | 4 | 0 | 3 |
| 7 | Recurrence<sup>2nd</sup> | 0 | 3 | 0 | 4 | 1 | 2 |
| 8 | Primary | 0 | 0 | 0 | 0 | 0 | 1 |
| 8 | Recurrence<sup>1st</sup> | 0 | 2 | 0 | 1 | 0 | 2 |
| 9 | Primary | 0 | 1 | 0 | 1 | 0 | 0 |
| 9 | Recurrence<sup>1st</sup> | 0 | 3 | 0 | 2 | 0 | 4 |
| 10 | Primary | 0 | 0 | 0 | 0 | 0 | 1 |
| 10 | Recurrence<sup>1st</sup> | 0 | 4 | 0 | 3 | 0 | 4 |
| 11 | Primary | 0 | 0 | 0 | 0 | 0 | 0 |
| 11 | Recurrence<sup>1st</sup> | 0 | 3 | 0 | 3 | 0 | 3 |
| 12 | Primary | 0 | 1 | 0 | 0 | 0 | 0 |
| 12 | Recurrence<sup>1st</sup> | 0 | 3 | 0 | 3 | 0 | 3 |
| 12 | Recurrence<sup>2nd</sup> | 0 | 2 | 0 | 4 | 0 | 3 |
| 13 | Primary | 0 | 0 | 0 | 0 | 0 | 1 |
| 13 | Recurrence<sup>1st</sup> | 0 | 2 | 0 | 1 | 0 | 2 |
| 13 | Recurrence<sup>2nd</sup> | 0 | 4 | 0 | 2 | 1 | 2 |
| 14 | Primary | 0 | 1 | 0 | 1 | 0 | 2 |
| 14 | Recurrence<sup>1st</sup> | 0 | 3 | 0 | 1 | 0 | 3 |
| 15 | Primary | 0 | 0 | 0 | 0 | 0 | 0 |
| 15 | Recurrence<sup>1st</sup> | 0 | 2 | 0 | 3 | 0 | 3 |
| 15 | Recurrence<sup>2nd</sup> | 0 | 3 | 0 | 4 | 0 | 3 |
| 16 | Primary | 0 | 0 | 0 | 0 | 0 | 0 |
| 16 | Recurrence<sup>1st</sup> | 0 | 2 | 0 | 2 | 1 | 3 |

**Table S3.** IHC score of VIM and MDK in Hypodermis.

| Type | VIM<br>(No of positive cell/area) | MDK<br>(No of positive cell/area) |
| --- | --- | --- |
| Normal Skin | 10 | 23 |
| Normal Skin | 7 | 42 |
| Normal Skin | 6 | 19 |
| Normal Skin | 13 | 50 |
| Normal Skin | 4 | 17 |
| Actinic Keratosis | 180 | 270 |
| Actinic Keratosis | 1052 | 505 |
| Actinic Keratosis | 430 | 230 |
| Actinic Keratosis | 588 | 698 |
| Actinic Keratosis | 273 | 1174 |
| Actinic Keratosis | 599 | 430 |
| Actinic Keratosis | 201 | 501 |
| Actinic Keratosis | 88 | 109 |
| Actinic Keratosis | 157 | 205 |
| Actinic Keratosis | 203 | 105 |

